# Modelling the effect of infection prevention and control measures on rate of *Mycobacterium tuberculosis* transmission to clinic attendees in primary health clinics in South Africa

**DOI:** 10.1101/2021.07.26.21260835

**Authors:** Nicky McCreesh, Aaron S Karat, Kathy Baisley, Karin Diaconu, Fiammetta Bozzani, Indira Govender, Peter Beckwith, Tom A Yates, Arminder K Deol, Rein MGJ Houben, Karina Kielmann, Richard G White, Alison D Grant

## Abstract

**Background:** Elevated rates of tuberculosis in health care workers demonstrate the high rate of *Mycobacterium tuberculosis (Mtb)* transmission in health facilities in high burden settings. In the context of a project taking a whole systems approach to tuberculosis infection prevention and control (IPC), we aimed to evaluate the potential impact of conventional and novel IPC measures on *Mtb* transmission to patients and other clinic attendees.

**Methods:** An individual-based model of patient movements through clinics, ventilation in waiting areas, and *Mtb* transmission was developed, and parameterised using empirical data from eight clinics in two provinces in South Africa. Seven interventions – co-developed with health professionals and policy-makers - were simulated: 1. queue management systems with outdoor waiting areas, 2. ultraviolet germicidal irradiation systems (UVGI), 3. appointment systems, 4. opening windows and doors, 5. surgical mask wearing by clinic attendees, 6. simple clinic retrofits, and 7. increased coverage of long antiretroviral therapy prescriptions and community medicine collection points through the CCMDD service.

**Results:** In the model, 1. outdoor waiting areas reduced the transmission to clinic attendees by 83% (interquartile range [IQR] 76-88%), 2. UVGI by 77% (IQR 64-85%), 3. appointment systems by 62% (IQR 45-75%), 4. opening windows and doors by 55% (IQR 25-72%), 5. masks by 47% (IQR 42-50%), 6. clinic retrofits by 45% (IQR 16-64%), and 7. increasing the coverage of CCMDD by 22% (IQR 12-32%).

**Conclusions:** The majority of the interventions achieved median reductions in the rate of transmission to clinic attendees of at least 45%, meaning that a range of highly effective intervention options are available, that can be tailored to the local context. Measures that are not traditionally considered to be IPC interventions, such as appointment systems, may be as effective as more traditional IPC measures, such as mask wearing.

## Introduction

All else being equal, the risk of tuberculosis from transmission in primary healthcare (PHC) clinics is likely to be higher than in many other types of congregate settings, due to higher rates of clinic attendance both by people with infectious tuberculosis, and by people at high risk of progression to disease^1^. Evidence for high rates of *Mycobacterium tuberculosis* (*Mtb*) transmission in health facilities can be found in studies of infection or disease risk in healthcare workers, with a recent systematic review finding an incidence of tuberculosis in healthcare workers in high burden countries 2–12 times higher than in the general population^2^.

The challenge of high rates of *Mtb* transmission in healthcare facilities comes with opportunities for control. Compared to many other putative high transmission risk congregate settings such as bars^3^ or public transport^4^, healthcare facilities should be relatively accessible settings for infection prevention and control (IPC) interventions. Two recent reviews, however, have identified a number of barriers to the successful implementation of IPC measures in healthcare facilities, including lengthy, ambiguous, or unclear guidelines; overwork; lack of training; lack of space and/or equipment; and concerns about patient stigmatisation^5,6^. The *Umoya omuhle* (‘good air’ in isiZulu) project was designed to address these barriers, taking a multidisciplinary whole systems approach to understanding the drivers of *Mtb* transmission in PHC clinics in South Africa, and the individual and system constraints to implementing IPC measures^7^. The project combined quantitative and qualitative data collection, and used a system dynamics modelling approach^8^ to identify potential IPC interventions. Interventions were selected that local policy makers and health professionals, working at PHC and province levels (including PHC and district level healthcare workers), ranked highly in terms of both feasibility of implementation and perceived likely impact^9^.

To make informed and evidence-based decisions on the implementation of IPC measures in PHC clinics, it is necessary to know the likely effects of the interventions on transmission risk. Empirical data on intervention impact are limited however, and focus on risk to healthcare workers, and on hospital settings^10^, likely due to the difficulties in empirically evaluating changes in risk to patients and other clinic attendees. We therefore use mathematical modelling to fill this key information gap, using a model of patient movement through clinics and ventilation rates (informed by empirical data on both) to estimate the potential effects of the interventions on the rate of *Mtb* transmission to clinic attendees in PHC clinics in KwaZulu-Natal and Western Cape provinces, South Africa. In doing so, we provide information that is critical to policy makers, to inform decisions on which intervention or interventions to implement in clinics.

## Methods

### Clinic attendee movement data

#### Collection

Clinic attendee movement data were collected on a single day per clinic, in six PHC clinics in KwaZulu-Natal province in February-March 2019, and five in Western Cape in May 2019 (with the exception of one clinic in KwaZulu-Natal, where data were collected on two separate days)^11^. Briefly, all patients and other clinic attendees (people attending with or on the behalf of patients) arriving at the clinic or present at the clinic at the start of data collection were given a unique barcode. Research staff with barcode scanners were positioned at key points throughout the clinic, including the facility entrance(s), the filing window where patients registered and collected their medical record, the triage/vitals station where measurements such as blood pressure were taken, and doorways of waiting areas and some consultation rooms. Each time that a clinic attendee passed through a doorway or visited a station (e.g. the filing window) their barcode was scanned, recording the time and the location. This allowed the attendees’ movements through the clinic to be tracked. Basic demographic information and information on visit reasons were collected from all attendees. Clinic staff were also assigned barcodes, and their movements tracked. Table 1 shows the number of attendees recorded at each clinic, the clinic opening time, and the time and number of attendees present at the start and end of data collection.

**Table 1.**
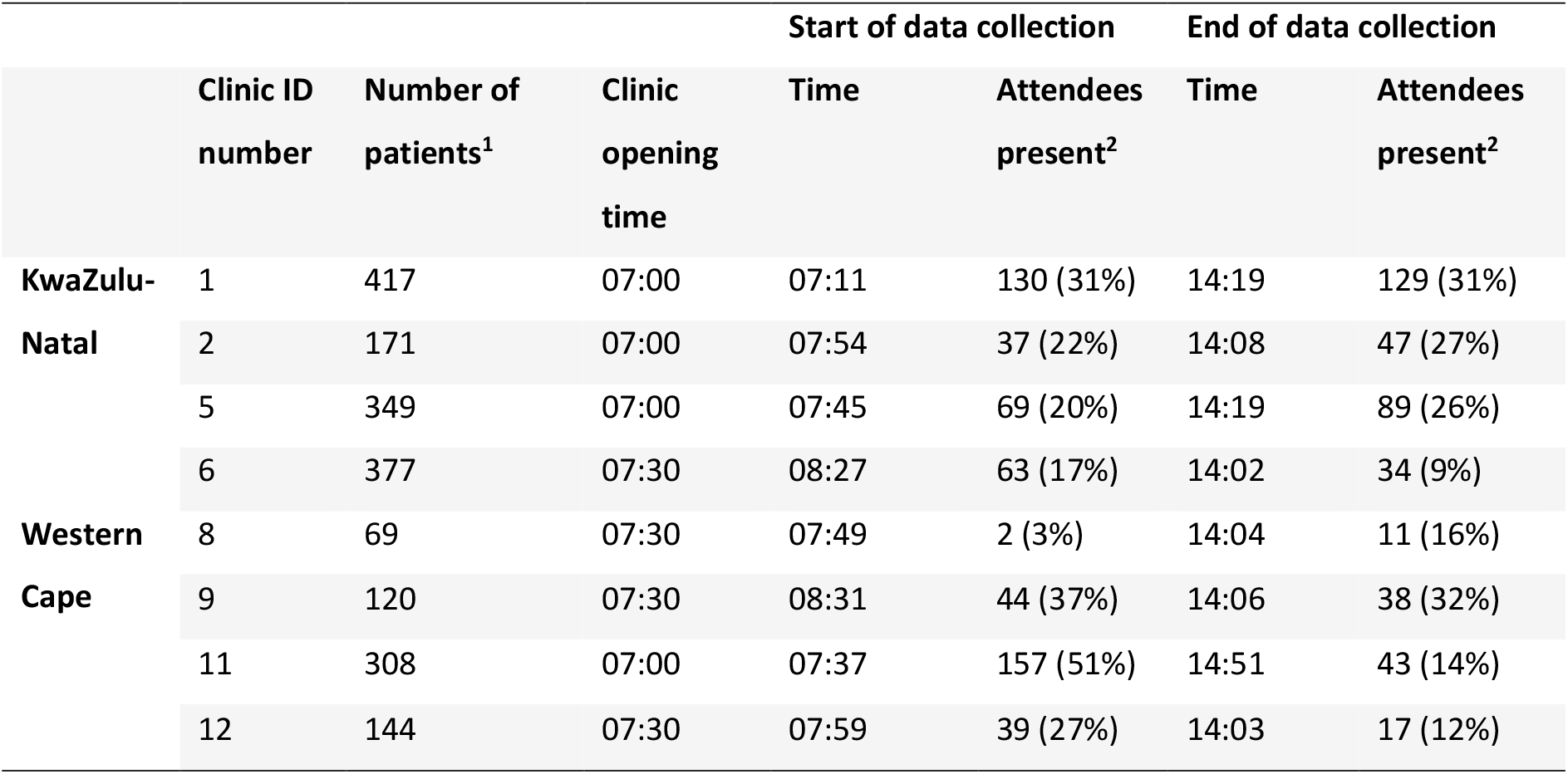
Clinic information. Clinic ID numbers correspond to numbers used in other papers from the *Umoya omuhle* project. ^1^Number of patients and other clinic attendees included in the data collection. ^**2**^Number and proportion of all patients and other clinic attendees included in the data collection who were already present at the start of data collection, or still present at the end.

Full details of the data collection methods and results are given in Karat *et al*^11^.

### Analysis

Data on clinic attendee movements were only collected from all main areas of the clinic in eight clinics, four in KwaZulu-Natal and four in Western Cape, and therefore only those clinics were considered in this work.

Attendees’ movements through the clinic were simplified to four key stages/times: the time they arrived at the clinic, the time that they were first recorded at the filing window (‘files’), the time that they were first recorded at the triage/vitals station (‘vitals’), and the time they left the clinic. All attendees were assumed to pass through each of the four stages, in order. These times were missing for many individuals, due to their barcode not being scanned, or their arrival or departure occurring outside the data collection period. Missing data on these times were imputed using multiple imputation (see supplementary material for details). The time that attendees started consultations was also estimated, from the attendees’ leaving times and data on mean consultation lengths.

Due to missing data on attendees’ movements between waiting areas, attendees’ waiting locations were also simplified, with each attendee assigned a single waiting area for each stage: waiting for files, waiting for vitals, and waiting for any consultations (including the pharmacy). Waiting areas were assigned based on recorded barcode scans into and out of waiting areas, and knowledge of clinic space use (see supplementary material for details).

In total, 40 baseline attendee datasets were created for each clinic, incorporating the uncertainty in the four times and three waiting locations.

### Ventilation and room size data

Data on ventilation rates were collected on 84 occasions from 57 rooms in 10 clinics in KwaZulu-Natal and Western Cape, using carbon dioxide (CO_2_) release experiments and continuous CO_2_ concentration data, with the room doors and windows in typical in-use configurations (‘usual conditions’). Room measurements were also made, and room volumes calculated. Ventilation rates, measured in air changes per hour (ACH), were calculated for each room. Full details are given in Beckwith *et al*^12^. and Deol *et al*^13^.

Data were also collected from 20 clinic rooms, with all windows and doors fully open (‘maximum ventilation conditions’). The relative change in ACH in maximum ventilation conditions was calculated for each room, relative to the ACH in usual conditions in the same room on the same day.

### Ethical approval

The clinic attendee movement and ventilation data collection received ethical approval from the Biomedical Research Ethics Committee of the University of KwaZulu-Natal (ref. BE082/18), the Human Research Ethics Committee of the Faculty of Health Sciences of the University of Cape Town (ref. 165/2018), the Research Ethics Committee of Queen Margaret University (ref. REP 0233), and the Observational/Interventions Research Ethics Committee of the London School of Hygiene & Tropical Medicine (ref. 14872).

### Model

#### Clinic attendee movements

We developed an individual-based model that tracked the time at each stage for each clinic attendee (arrival, files, vitals, and leaving), and the locations that they were waiting in between stages. The estimated mean number of quanta (‘infectious doses’^14^) in each waiting area of the clinic was tracked over time, assuming that 1% of adult patients^15^ and 0.02% of child patients^15–17^ had potentially infectious tuberculosis. Estimates of ventilation rates were used to determine the rate at which quanta were cleared from the room. Cumulative infection risk over time was tracked both for each individual attendee, and by room. Full details of the model and model parameterisation are given in the supplementary material.

**Figure 1.**
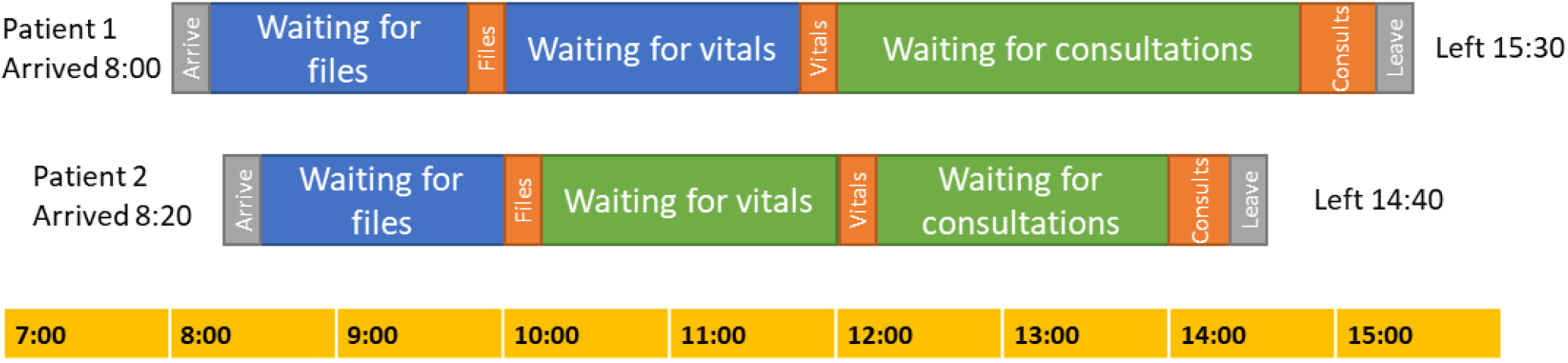
Example illustration of the movement of two hypothetical patients through a clinic in the model. The blue and green shading indicate different waiting areas.

### Interventions

Seven potential IPC interventions had been identified through qualitative research and system dynamics modelling workshops conducted as part of the *Umoya omuhle* project^9^. They were implemented in the model as follows:

1. **Opening windows & doors**. Ensuring windows and doors in waiting areas are kept open at all times. This was implemented in the model through increased ACHs, with the relative increase in each waiting area and model run sampled from a distribution fitted to the empirical ventilation data.
2. **Simple clinic retrofits**. Retrofits are changes to the building to improve ventilation rates. This could include installing lattice brickwork or whirlybird fans. Due to the large amount of variation between clinic spaces in the types of building retrofits that would be suitable, and the lack of sufficient data on the effects of the retrofits on ventilation rates in different types of spaces, we did not model specific retrofits or packages of retrofits. Instead, we simulated an undefined package of retrofits that are sufficient to increase air changes per hour to a minimum of 12 in all rooms, chosen in line with WHO guidelines^2,18^
3. **UVGI systems**. We assume in this intervention that appropriate and well-maintained ultraviolet germicidal irradiation (UVGI) systems are installed in all indoor clinic waiting areas. This was implemented in the model through an additional quanta clearance rate, equivalent to a ventilation rate of 24 ACH (95% CI 9.9–62)^19^.
4. **Surgical mask wearing by clinic attendees**. Based on discussions with health care workers and professionals active in the management of health services in the two provinces we worked in, as well as review of qualitative data collected, we determined that a scenario where 70% of attendees wear surgical masks 90% of the time was plausible. This was implemented in the model as 63% of attendees wearing masks 100% of the time, with the attendees who wear the masks chosen at random each model run. Masks were assumed to reduce the rate of quanta production by 75% (95% CI 56– 85%)^20^, and have no effect on rate of infection for the person wearing the mask^21^.
5. **Increased CCMDD coverage**. South Africa’s Central Chronic Medicine Dispensing and Distribution (CCMDD) programme is designed to allow patients with stable chronic health conditions to collect their medicines from convenient locations, such as local pharmacies^22^. This means that they do not need to queue at clinics unnecessarily. The purpose of this intervention is to increase the coverage of CCMDD and similar programmes for eligible patients on antiretroviral therapy (ART), and to ensure that pick-up points do not require patients to queue at clinics. We assumed that 92% (95% CI 84–95%) of patients could have their ART appointments reduced to once every 6 months^23^, and that the remaining 8% of people need monthly ART appointments. This was implemented in the model through removing 31% (interquartile range [IQR] 22– 34%) of patients attending for HIV care, chosen at random each model run.
6. **Queue management system with outdoor waiting areas**. Empirical data show that clinic waiting areas are often crowded, and that in many clinics patients wait in unsuitable areas such as corridors^11^. Conversations with clinic staff suggested that this is partly due to patient concerns that if they wait in other areas, they may not hear their name being called, and may miss their turn. This intervention therefore combines a large, covered outdoor waiting area with a queue management system. We assumed that only 5–10 patients would be allowed to wait inside the clinic for each of the three stages, with the rest waiting in a large, covered, outdoor waiting area, with a very high ventilation rate of 52–70 ACH^24^.
7. **Appointmdnt system**. In this intervention, we simulated an appointment system to reduce clinic overcrowding, through spacing out the arrival times of patients. As date-time appointment systems were already in place in some form in the Western Cape clinics on the day that the patient data were collected, we only modelled the appointment intervention in the KwaZulu-Natal clinics. We assumed that all patients aged <16 years and a proportion of patients with acute visit reasons would arrive at the clinic at the same time as in the baseline scenario, and be seen the same day. We assumed that all adult chronic patients, and a proportion of adult acute patients would be given appointments, with their arrival time spaced out between 9am and 2pm.

The CCMDD intervention reduces the number of patients, and the appointment system intervention changes the arrival times of some patients. Both these interventions may have consequences for the times that other patients are seen at each stage (files, vitals, and consultations/leaving). The consequences are likely to vary by stage, and will vary depending on whether or not the stage is rate limiting. In other words, does the stage usually have the capacity to see patients as soon as they are ready, or are there usually queues? The model therefore contained two scheduling mechanism options per stage, which assume that the stage is or is not rate limiting.

Full details of the model and simulated interventions are given in the supplementary material.

### Model runs and uncertainty estimation

For each of the 40 patient datasets (incorporating the uncertainty in the times and waiting locations), 100 ventilation input sets were created, with the baseline ACH in each room varying between input sets. For each of the 4000 combinations, four different scheduling scenarios were simulated, assuming that the files and vitals stages are or are not rate limiting, in a two-by-two factorial design. Consultations/leaving was assumed to be a rate limiting stage in all the main model runs. This gave a total of 16,000 model runs for each clinic and intervention.

In addition to this, as a sensitivity analysis, an additional 16,000 model runs were done for each clinic and intervention, where it was assumed that consultations were not a rate limiting stage.

## Results

### Effect of the interventions on the relative rate of transmission to patients

Figure 2 shows the estimated reduction in the rate of *Mtb* transmission to patients in each of the intervention scenarios, compared to the baseline scenario, overall and by province. Overall, in the model, opening windows and doors reduced the transmission rate by 55% (IQR 25%–72%), clinic retrofits by 45% (IQR 16%–64%), installing UVGI by 77% (IQR 64%–85%), surgical mask wearing by patients by 47% (IQR 42%–50%), increasing the coverage of CCMDD by 22% (IQR 12%–32%), and a queue management system plus outdoor waiting area by 83% (IQR 76%–88%). In the KwaZulu-Natal clinics, implementing an appointment system in the model reduced the transmission rate by 62% (IQR 45%–75%).

**Figure 2.**
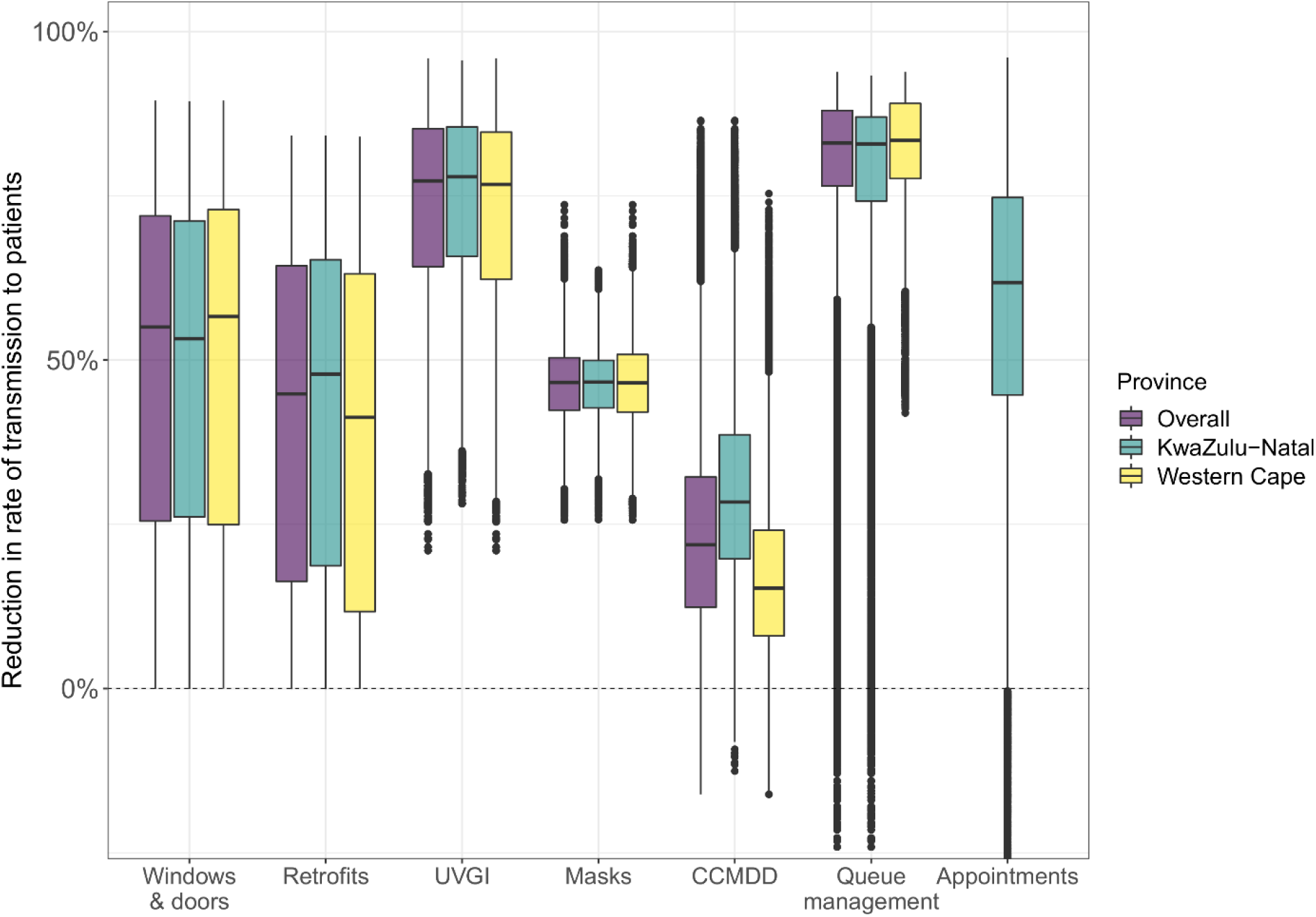
Estimated reduction in the rate of *Mycobacterium tuberculosis* transmission to patients in clinics, by province and intervention. The central line indicates the median, the box range the interquartile range (IQR), the whiskers the most extreme value within 1.5 * IQR from the box, and the points outlying values. In the queue management intervention in KwaZulu-Natal, 1.3% of points were below −20%, with a minimum of −162%. In the appointment system intervention in KwaZulu-Natal, 1.3% of points were below −20%, with a minimum of −83%. These points are not shown on the graph. The appointment system intervention was not modelled in Western Cape, due to the presence of existing appointment systems. UVGI stands for ultraviolet germicidal irradiation, and CCMDD for Central Chronic Medicine Dispensing and Distribution.

There was little variation in estimated impact by province, with the exception of increasing the coverage of CCMDD, where reductions in the transmission rate were higher in KwaZulu-Natal clinics (28% IQR 20%–39%) than in Western Cape clinics (15% IQR 8%–24%), reflecting the higher prevalence of HIV and higher ART coverage in KwaZulu-Natal.

Figure 3 shows the number of patients in the clinic 1 over time in the baseline scenario, then with the appointment system, and the CCMDD intervention. The lower panels show the mean rate of transmission to each patient in the clinic over time in all scenarios. Similar figures for the other clinics are in the supplementary material.

**Figure 3.**
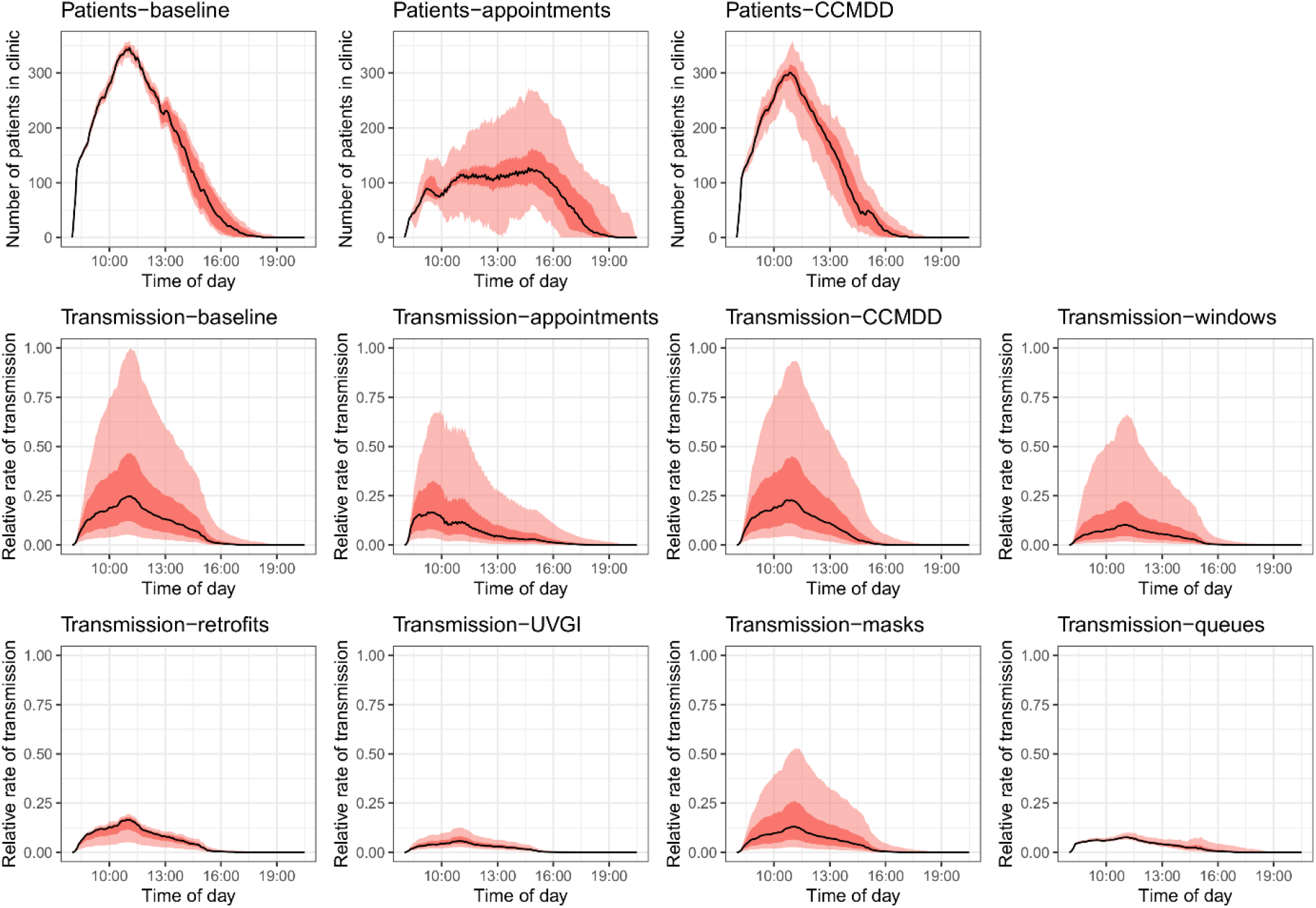
Number of patients in the clinic over time in the baseline, appointments, and CCMDD interventions, and the mean rate of transmission to each patient in the clinic over time in all scenarios, for clinic 1. The black line shows the median result, the dark red band the interquartile range, and the light red band the 95% plausible range. For interventions where a plot of the number of patients over time is not shown, the intervention has no effect on patient numbers. Transmission rates are relative to the highest transmission rate in any scenario at any point in time. UVGI stands for ultraviolet germicidal irradiation, and CCMDD for Central Chronic Medicine Dispensing and Distribution. Figures for the other clinics are shown in the supplementary material.

### Sensitivity analyses

Simulating consultations as a non-rate limiting stage reduced the estimated reduction in the rate of transmission from 22% (IQR 12%–32%) to 15% (IQR 8.7%–23%) in the CCMDD scale-up intervention, and from 62% (IQR 45%–75%) to 24% (IQR 13%–47%) in the appointments intervention (Figure S3).

## Discussion

In this paper, we estimated the potential effects of seven interventions on the rate of *Mtb* transmission to patients and other clinic attendees in PHC clinics in two provinces in South Africa. A queue management system with outdoor waiting areas and installing UVGI systems were identified as the most effective interventions, reducing the rate of transmission by an estimated 83% and 77% respectively. The majority of interventions resulted in substantial reductions in the transmission rate however, demonstrating that a range of highly effective IPC measures exist. This includes appointment systems, which are not traditionally considered as IPC measures. This highlights the benefits of broadening our views of IPC and expanding our view of the population to be protected beyond healthcare workers, to also include patients and other clinic attendees.

Many of the interventions could be implemented in different ways in practice, increasing or decreasing their effects. For example, installing a more extensive package of retrofits, or taking measures to achieve a higher level of mask wearing. The COVID-19 pandemic may also have led to changes in the way that PHC clinics operate, some of which may last beyond the end of the epidemic. For example, the acceptability to clinic attendees of mask wearing may increase, increasing the coverage that can be achieved. When interpreting the results, consideration should therefore be given to any differences in the way that we implemented the intervention in the model, and the way that they would be implemented in a specific context. Nevertheless, our results provide a useful baseline estimate of the impacts and relative impacts of the different interventions.

The choice of IPC intervention(s) to implement at the clinic level, or to recommend at a district or provincial level, will necessarily also be guided by other factors. The costs of implementing and maintaining the different interventions will be a key factor, and is being explored in further work as part of the *Umoya omuhle* project. Guided by a whole systems approach, we have comprehensively costed the interventions proposed by also considering how to overcome potential system and practical barriers to implementation. The ease and practicality of implementation is also an important consideration, and will vary by clinic. For instance, is there sufficient space to install an outdoor waiting area? And is the climate suitable for interventions that increase natural ventilation rates? The systems dynamics modelling work conducted to identify the interventions simulated here also aimed to identify the mechanisms necessary to achieve the interventions. For instance, for ensuring an effective queue management programme, mechanisms such as community and health service staff consultation and creation of covered outdoor waiting areas were discussed^9^.

Some of the interventions have additional benefits to patients. Improving the coverage of CCMDD may be beneficial to patients stable on ART, reducing the amount of time they spend queuing at clinics, and the financial cost to patients. An appointment system should also reduce the time spent at clinics for the majority of patients. The effect of the interventions on risk of transmission to healthcare workers and other clinic staff should also be considered. All the interventions described here will reduce risk to all staff situated in waiting areas, such as security guards and clerks in some clinics. Many of the interventions will also reduce risk to staff in consultation rooms, provided that the interventions are also implemented in those spaces. Interventions that reduce risk by reducing overcrowding in waiting areas (appointment systems, CCMDD scale-up, and outdoor waiting areas) will have little effect on risk for staff based in consultation rooms however.

Finally, we estimate the effect of the interventions on an airborne infection, *Mycobacterium tuberculosis*. The relative effects of the different interventions on other infections that spread primarily through airborne transmission, such as measles and chickenpox, are likely to be similar, although the concentration of these infections in children rather than adults may alter the effects slightly. The effects will differ, however, for infections where droplet or fomite transmission plays a larger role, such as SARS-CoV-2 and influenza. Fully exploring the impact on these infections is beyond the scope of this paper, however, as a rough guide, interventions that act through reducing patient concentrations (CCMDD and appointments systems) or reducing the release of pathogens (masks) will have a greater effect on these infections than interventions that act through improving ventilation (opening windows and doors, clinic retrofits, outdoor waiting areas) or air disinfection (UVGI).

There are a number of limitations to this work. Firstly, empirical data on the flow of clinic attendees through clinics were only available for one day per clinic, meaning that we cannot disentangle variation between the simulated clinics in the intervention effects that arises from day-to-day variation within the clinic from that which arises from variation between clinics. For this reason, the results are presented by province only in the main results figure, rather than by clinic. Additional empirical data (both on patient movements and ventilation rates in waiting areas), and simulated clinic days, would also increase the confidence that our results incorporate the full range of variation between clinics and clinic days. Similarly, data from additional clinics would increase the generalisability of our results.

Secondly, there were large amounts of missing data in the clinic attendee movement datasets. Missing data were imputed using multiple imputation, and the effects of the uncertainties in patient times and waiting locations were reflected in the size of the uncertainty bounds around the results. Multiple imputation relies on the assumption that the data are missing at random however, which may not be true for our datasets. We also assume that all clinic attendees visited both files and vitals in turn, and that all attendees waited in a single location per stage, which may not have been the case for all attendees. For these reasons, our results for each clinic day should be considered to be indicative of the interventions’ effects in the clinic, rather than a definitive estimate of the effects of the interventions on each specific clinic day. We may also have missed a number of attendees entirely if they left before the start of data collection, or arrived after the end. The effects of this are likely to have been minimal however, as attendees leaving before the start of data collection could only have spent a small amount of time at the clinic, and attendee numbers were relatively small and fell rapidly after the end of data collection^11^.

Due to the large amounts of missing data in the clinic attendee movement datasets, we were also unable to simulate in any detail the process of queuing for consultations with nurses and doctors, and for the pharmacy. Instead, we simulated a single queue for consultations, using data on clinic leaving times. This is unlikely to have had a substantial effect on the estimates for the majority of interventions, but may mean that we under- or over-estimated the effects of the appointment systems and CCMDD coverage scale-up interventions. The large amounts of missing data also prevented us from considering the pathways of people who were accompanying patients or attending on the behalf of someone else separately from the pathways of patients, and all clinic attendees are treated as ‘patients’ in the model. This is unlikely to have had a large effect on the results, as the people accompanying patients are likely to have spent the majority of the time in the same waiting areas as the patients they were accompanying.

In a small proportion of runs, the simulated interventions increased the rate of transmission. In the CCMDD and appointment interventions, this occurred through rearrangements in patient pathways making waiting times for some patients higher, or concentrating patients in higher risk waiting areas. These rare outliers reflect day-to-day variation, rather than highlighting a real potential for the interventions to consistently increase risk. For the queue management intervention, increased rates of transmission occurred in model runs where high sampled baseline ventilation rates in waiting areas coincided with a low sampled ventilation rate in the outdoor waiting area in the intervention scenario. In reality, ventilation rates in different areas will be correlated, with both influenced by the same factors such as wind speed^25^, and it is therefore likely that the outlier runs overestimate the true uncertainty.

## Conclusions

To conclude, we show the estimated effects on the rate of *Mtb* transmission to clinic attendees of a range of IPC infections. Median reductions range from 83% for a queue management system with outdoor waiting areas, to 22% from scaling up coverage of CCMDD among ART patients. The majority of the interventions (6/7) achieve reductions of at least 45%, meaning that a range of highly effective conventional and novel IPC intervention options are available, that can be tailored to the local context.

## Data Availability

The mathematical model used in this work is available from https://github.com/NickyMcC/WithinClinics The datasets used will be made available from https://datacompass.lshtm.ac.uk/ on publication of the manuscript.

https://github.com/NickyMcC/WithinClinics

## Funding/acknowledgements

The support of the Economic and Social Research Council (IK) is gratefully acknowledged. The project is partly funded by the Antimicrobial Resistance Cross Council Initiative supported by the seven research councils in partnership with other funders including support from the GCRF. Grant reference: ES/P008011/1. NM is additionally funded the Wellcome Trust (218261/Z/19/Z). RGW is funded by the Wellcome Trust (218261/Z/19/Z), NIH (1R01AI147321-01), EDTCP (RIA208D-2505B), UK MRC (CCF17-7779 via SET Bloomsbury), ESRC (ES/P008011/1), BMGF (OPP1084276, OPP1135288 & INV-001754), and the WHO (2020/985800-0). TAY is funded via an NIHR Academic Clinical Fellowship. RMGHJ is funded by ERC (action number 757699)

We would like to thank the managers, staff, patients, and visitors at participating clinics, and all of the Umoya omuhle project team involved in the data collection. Full details are given in the supporting material.

