## Supplementary material for "Modelling the effect of infection prevention and control measures on rate of *Mycobacterium tuberculosis* transmission to clinic attendees in primary health clinics in South Africa"

1.10.3 UVGI systems ... 17

1.10.4 Surgical masks wearing by clinic attendees ... 17

1.10.5 Increased CCMDD coverage ... 18

1.10.6 Queue management system and outside waiting areas ... 20

1.10.7 Appointment systems ... 21

1.11 Input parameter values ... 23

2 Supplementary results ... 26

2.1 Sensitivity analysis ... 26

2.2 Intervention impact by clinic ... 27

2.3 Attendee numbers and rate of transmission over time ... 28

2.4 Median clinic visit durations ... 35

**3** Supplementary acknowledgements ... 35

4 References ... 38

### 1 Supplementary methods

#### 1.1 Imputation

##### 1.1.1 Times

Four key times were identified in the pathways that each clinic attendee took through the clinic:

- Arrival time. The time that they first arrived at the clinic. For attendees who arrived after the start of data collection, this was assumed to be the time that their barcode was first scanned. The arrival time was set to missing if the attendee was already present in the clinic before the start of data collection, or if the first time their barcode was scanned was not at a clinic entrance (an external door or compound gate).
- Files time. The time that the attendee obtained their patient file from the clinic reception desk. This was assumed to be the time that their barcode was first scanned at files, provided that it occurred before the first time that they were scanned at vitals or at a consultation room. The time was set to missing if they never scanned at files, or if they scanned at vitals or a consultation room before first scanning at files.
- Vitals time. The time that the attendee has their blood pressure, heart rate, and respiratory rate measured. This was assumed to be the time that their barcode was first scanned at vitals, provided that it occurred before the first time that they were scanned at a consultation room. The time was set to missing if they never scanned at vitals, or if they scanned at a consultation room before first scanning at vitals.
- Leave time. The time that the attendee left the clinic. This was assumed to have occurred at the final time that they scanned their barcode, provided it occurred at a clinic exit point (an external door or compound gate). The leaving time was set to missing for attendees who were still at the clinic at the end of data collection, or if their barcode was never scanned at an exit point.

In a small number of cases, times at files and/or vitals may be missing not because the attendee did not scan their barcode, but because the attendee did not complete that stage. For instance, some attendees who were at the clinic to collect medicine only may have skipped one or both stages. In many clinics, patients on TB treatment can also skip the files and vitals stages. In all eight clinics however, the majority of patients are required to pass through both files and vitals, regardless of their visit reason.

Table S1 shows the number and proportion of attendees with known and missing data for each stage.

| | | | | Arrival | | | Files | | Vitals | | Leaving | | |
|---|---|---|---|---|---|---|---|---|---|---|---|---|---|
| | Clinic | Clinic closing time | Number of attendees | Known | Missing (arrived early[1]) | Missing | Known | Missing | Known | Missing | Known | Missing (left late[2]) | Missing |
| **KwaZulu-Natal** | 1 | 19:00 | 417 | 269 (65%) | 130 (31%) | 18 (4%) | 66 (16%) | 351 (84%) | 34 (8%) | 383 (92%) | 248 (59%) | 129 (31%) | 40 (10%) |
| | 2 | 17:00 | 171 | 130 (76%) | 37 (22%) | 4 (2%) | 62 (36%) | 109 (64%) | 66 (39%) | 105 (61%) | 121 (71%) | 47 (27%) | 3 (2%) |
| | 5 | 16:30 | 349 | 257 (74%) | 69 (20%) | 23 (7%) | 14 (4%) | 335 (96%) | 38 (11%) | 311 (89%) | 247 (71%) | 89 (26%) | 13 (4%) |
| | 6 | 17:00 | 377 | 128 (34%) | 63 (17%) | 186 (49%) | 99 (26%) | 278 (74%) | 109 (29%) | 268 (71%) | 174 (46%) | 34 (9%) | 169 (45%) |
| **Western Cape** | 8 | 16:30 | 69 | 65 (94%) | 2 (3%) | 2 (3%) | 35 (51%) | 34 (49%) | 23 (33%) | 46 (67%) | 55 (80%) | 11 (16%) | 3 (4%) |
| | 9 | 16:30 | 120 | 56 (47%) | 44 (37%) | 20 (17%) | 34 (28%) | 86 (72%) | 40 (33%) | 80 (67%) | 54 (45%) | 38 (32%) | 28 (23%) |
| | 11 | 16:30 | 308 | 111 (36%) | 157 (51%) | 40 (13%) | 32 (10%) | 276 (90%) | 24 (8%) | 284 (92%) | 176 (57%) | 43 (14%) | 89 (29%) |
| | 12 | 16:30 | 144 | 94 (65%) | 39 (27%) | 11 (8%) | 39 (27%) | 105 (73%) | 66 (46%) | 78 (54%) | 121 (84%) | 17 (12%) | 6 (4%) |

**Table S1. The number and proportion of attendees with known and missing data for each stage, and clinic closing times** [1]The person arrived before the start of data collection. [2]The person left after the end of data collection

Missing times were imputed as interval-censored values, with lower and upper bounds of when the event would have occurred, using a sequential approach. Firstly, arrival times at the clinic were multiply-imputed using 20 imputations. For attendees who arrived before the start of data collection, the lower and upper limits of the time of arrival were set to the clinic opening time and the start of data collection, respectively. For those who arrived after the start, the lower limit was set as the start of data collection, and the upper limit was the time that the attendee was first scanned. Secondly, the time at files was imputed, using the imputed arrival time as the lower bound of the interval and time at vitals (if observed) as the upper limit. If time at vitals was not observed, the upper bound was set to the earliest of the maximum time from arrival to files observed in that clinic, the time of leaving (if observed), end of data collection (if not there at end) or close of clinic (if there at end). Next, the time at vitals was imputed, using the imputed time at files as the lower bound of the interval, and the setting the upper bound to the earliest of the maximum time from files to vitals observed in that clinic, the time of leaving (if observed), end of data collection (if not there at end) or close of clinic (if there at end). Finally, the time of leaving the clinic was imputed, using the imputed time at vitals as the lower bound, and setting the upper bound to the earliest of the maximum time from vitals to leaving observed in that clinic, end of data collection (if not there at end) or close of clinic (if there at end).

Age, sex, clinic, reason for visit, whether there at start/end, and whether the attendee was first scanned in the morning (before 10am) were included in the imputation model. Two sets of 20 imputations were generated. In one, separate lower and upper limits were used for the morning and afternoon visits. This was done as there was some evidence in the empirical data that waiting times were shorter in the afternoons. In the second, the same lower and upper limits were used for all attendees.

For each attendee and imputation, an estimated time at consultations was generated. This was not designed to be an accurate estimate of the exact time that they started any particular consultation, but instead was used to ensure that the time that attendees spent in waiting areas between vitals and leaving the clinic was not over-estimated. Observations in clinics suggested a mean time per consultation of seven minutes. We assumed that patients have an average of 1.5 consultations per visit, giving a mean length of time spent in consultations of 10.5 minutes. We also assumed that the majority of patients would need a minimum of 3 minutes between starting vitals and starting their first consultation. Finally, the estimated time starting consultations, 'consultation time', could not occur after 'leave time'. The estimated consultation time was therefore set to the latest of 1) attendees leave time – 10.5 minutes, 2) vitals time + 3 minutes, 3) leave time.

The files and vitals stages only take a short amount of time per patient, and in many clinics the patient remains in the files waiting area while their file is being retrieved. The time not spent in the waiting area for files and vitals is therefore considered to be negligible, and is not subtracted from the patients' waiting times in the model.

##### 1.1.2 Locations

We assume that each attendee waits in a single location for each stage of their clinic pathway. That is:

- Between arrival time and files time
- Between files time and vitals time
- Between vitals time and consultation time

Based on observation of the organisation of care and patient flow at each clinic, each stage was assigned a certain area or areas in which individuals would wait to be seen. For each individual, waiting locations for each stage were determined in three steps.

1. For individuals who had a recorded visit to a specific stage:

a. The location recorded immediately before the stage was used as the most likely waiting location if it was one of the waiting areas associated with that stage.

b. For stages with only one associated waiting location, individuals who had a recorded visit to a particular stage, but whose immediate previous location was not the waiting area for that stage, were nevertheless listed as having waited in that area, as it was considered likely that their entry and exit to that waiting area had been missed during data collection. For stages with more than one waiting location, individuals were randomised to one of the areas using the method described in point 3 below.

c. For individuals who visited more than one consultation room, the first consultation room visited and associated waiting area were used.

2. Individuals without a recorded visit to a specific stage (any of filing, vitals, or consultation) were assigned waiting locations based on the organisation of care at the clinic.

    a. In clinics with a single filing and/or vitals stage, and where that stage had only one associated waiting area, all individuals were listed as having waited in the associated waiting area for a particular service. In clinics where a stage had more than one waiting area, individuals were randomised as described below.

    b. In clinics with more than one filing and/or vitals stages (e.g., clinics with separate streams for 'acute' and 'chronic' patients), individuals were first categorised by stream, based on the reported reason for their visit and by the consultation room they had attended (if recorded). Once again, if a stage had only one associated waiting area (e.g., 'acute vitals'), all individuals in the appropriate stream (e.g., the 'acute' stream) were listed as having waited in that area. If a stage had more than one associated waiting area, individuals were randomised as described below.

3. After the completion of steps 1 and 2, any individuals without recorded waiting locations for any of the three stages were assigned at random to a waiting area associated with that

stage. For each stage, the proportions of individuals to be assigned to each associated waiting areas was calculated using the assignments made in steps 1 and 2 above. The remaining individuals were then randomised to the associated waiting areas in the same proportions

A total of 20 attendee waiting location datasets were created for each clinic, incorporating the uncertainty in waiting locations.

The numbers and proportions of attendees with uncertain waiting locations (separated by waiting locations assigned by high probability [step 2, above] and by randomisation [step 3]) are shown in Table S2.

| | | | | Number of attendees with uncertain waiting location†, n (%) | | | | | |
|---|---|---|---|---|---|---|---|---|---|
| **Province** | **Clinic** | **Number of attendees** | **Number of waiting areas*** | **Files** | | **Vitals** | | **Consultations** | |
| | | | | **High probability** | **Randomised** | **High probability** | **Randomised** | **High probability** | **Randomised** |
| **KwaZulu-Natal** | 1 | 417 | 2 | 92 (22) | 171 (41) | 113 (27) | 140 (33) | 111 (26) | 196 (47) |
| | 2 | 171 | 4 | 0 | 107 (63) | 0 | 89 (52) | 0 | 83 (49) |
| | 5 | 349 | 4 | 142 (41) | 183 (52) | 290 (83) | 0 | 190 (54) | 98 (28) |
| | 6 | 377 | 4 | 268 (71) | 0 | 257 (68) | 0 | 172 (45) | 60 (16) |
| **Western Cape** | 8 | 69 | 2 | 15 (22) | 0 | 34 (49) | 0 | 25 (36) | 0 |
| | 9 | 120 | 2 | 50 (42) | 0 | 59 (49) | 0 | 60 (50) | 0 |
| | 11 | 308 | 5 | 275 (89) | 0 | 281 (91) | 0 | 295 (96) | 0 |
| | 12 | 144 | 2 | 78 (54) | 0 | 61 (42) | 0 | 11 (7.6) | 111 (77) |

**Table S2. The numbers and proportions of attendees with uncertain waiting locations, by clinic and stage.** *includes informal waiting areas such as corridors. Informal and formal outdoor waiting areas are not included in this number. †'Uncertainty' separated into those to whom waiting location was assigned based on high probability (step 2 in text) or randomisation (step 3)

#### 1.2 Ventilation data

Empirical data on air changes per hour (ACH) were available from a series of experiments conducted in a range of different rooms in the clinics[1,2]. Results from a small number of repeat experiments in the same rooms on different days showed that there were large amounts of variation in ventilation rates in the same room between different days. Estimated ACH were available from 84 experiments in 57 rooms (33 experiments in consultation rooms, and 51 in waiting areas) with a typical in-use number of windows and doors open or closed. There was little difference between estimated ACH between consultation rooms and waiting areas (mean 13.1 95% CI 8.2-18.0 and 18.5 95% CI 12.6-24.4, p=0.2), and therefore data from both types of area were used. An exponential distribution was fitted to the estimated rates (Figure 1), and simulated rates in each room were sampled from the distribution for each model run. Estimated air change rates per hour were similar between the data (mean 16.4, median 9.5, IQR 4.3-21.2) and the modelled distribution (mean 15.5, median 9.8, IQR 4.1-23.4).

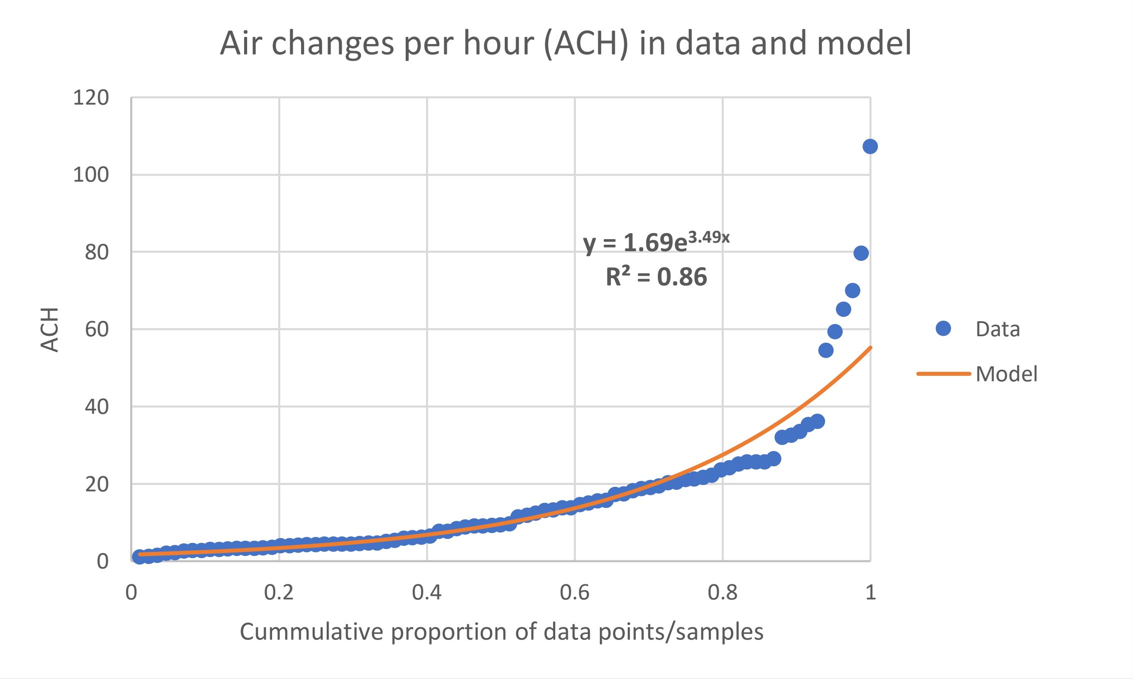

**Figure S1. Empirical data on air changes per hour (ACH), and distribution used to generate ACH values in the model**

#### 1.3 Model overview

The model was an individual-based model that tracks the movements of attendees through clinics, and *Mycobacterium tuberculosis* infection risk in clinic waiting areas over time, by area and by individual.

In the model, four key (clock) times control each attendee's movement through the clinic, through four corresponding stages: the time they arrive at the clinic, the time they collect their patient file ('files'), the time that their basic measurements are taken ('vitals'), and the time that they start consultations. It is assumed that they leave the clinic immediately after ending consultations, and spend negligible further time in waiting areas. Simulated attendees also each have an assigned waiting area where they wait between each stage (between arrival and files, between files and vitals, and between vitals and consultations). The four key times and three locations were determined, imputed and/or estimated for each attendee, and the complete dataset was used as input to the model. The simulated times and waiting locations remain unchanged in the model from those in the input files, in the baseline scenario and the majority of the intervention scenarios. In the appointment system and CCMDD interventions, the times are changed in the model, and in the queue management system, the waiting locations are changed.

The number of quanta in each waiting area is tracked over time. It is assumed that there is a prevalence of pulmonary tuberculosis among adult and child attendees of 1.0% and 0.016%[3-5] respectively, and that attendees with pulmonary tuberculosis have a mean rate of quanta production of 1.25 per hour[6]. We implement this in the model by giving each adult and child a rate of quanta production of $8.9 \times 10^{-3}$ and $1.42 \times 10^{-4}$ per hour respectively.

#### 1.4 Key

Model parameter names are written in *italics*, with colour indicating whether the parameter is an input parameter, a parameter with a global model-wide value, calculated from input parameter(s);or an individual-level parameter, which can take a different value for each simulated person, for each simulated waiting area, or for each simulated stage.

#### 1.5 Attendee input file

Each model run for each clinic required an attendee input file, which had a row for each attendee, with the following information:

1. The attendee id
2. The arrival time to be simulated for the attendee (equal to the imputed arrival time for all scenarios except the appointments intervention) (*arrival_time*)
3. The gap in the imputed data between the time they start a stage and the time that the next attendee starts the stage, for each stage (*duration_files*, *duration_vitals*, *duration_cons*)
4. The gap in the imputed data between each of their own stages (*gap_files*, *gap_vitals*, *gap_cons*)
5. The waiting location for each stage (*files_queue_location*, *vitals_queue_location*, *cons_queue_location*)

Each attendee input file contained the same number of attendees for each clinic, with the exception of input files for the CCMDD intervention, where a proportion of attendees were removed (see section 'intervention scenarios'). The gaps between attendees in the input file (*duration_files*, *duration_vitals*, and *duration_cons*) were not affected by the removal of attendees.

#### 1.6 Movement through clinics

All attendees enter the clinic at *arrival_time*, and set their location to *files_queue_location* (with the exception of the queue management intervention-see Attendee waiting areas). The way that the movement through the other three stages (files, vitals, and consultations) is implemented in the model – the scheduling mechanism – depends on whether the stage is set to be rate limiting or not, for that particular model run. In practice, whether stages are set to be rate limiting or not has no effect on model output for the baseline scenario, or for the majority of intervention scenarios, as in both cases the scheduling mechanisms result in the simulated times at which attendees reach each stage being exactly equal to the corresponding times in the attendee input file. The choice of scheduling mechanism for each stage only effects the results when the number of attendees are changed (CCMDD intervention), or attendee arrival times are changes (appointment systems).

Observations in the clinics suggested that the consultations stage was rate limiting for the majority of patients, with patients queueing for consultations throughout the day. Consultations were therefore assumed to always be rate limiting in the main model runs.

It was not possible to determine whether the files and vital stages were rate limiting in the eight clinics on the day of data collection, due to the large amounts of missing data. Whether a stage is rate limiting or not may also vary over the course of a day. For instance, the files stage may potentially be rate limiting at the start of the clinic day only. Which stages are rate limiting is also to some extent a function of staff allocation. Blockages at files and vitals in particular can be alleviated, through assigning additional staff to those stages. That may not be possible for consultation stages however, where more specific staff skills may be required. For these reasons, we simulated four scheduling scenarios, with both files and vitals simulated as rate limiting, with neither simulated as rate limiting, and with only one simulated as rate limiting.

##### 1.6.1 Scheduling mechanism – rate limiting stages

When the files stage is set to be rate limiting in the model, then the gap between each attendee and the attendee after them (*gap_files*) is kept the same as it is in the attendee input file. The files stage has a variable, *files_status*, that tracks whether there is somebody currently at the stage ('busy'), or whether there is not ('free'). At the start of the model run, *files_status* is set to free.

When *files_status* is set to free, then the next attendee to arrive at the clinic (i.e. finish the preceding stage) immediately starts the files stage, setting *files_status* to 'busy'. When they finish the files stage, after a gap of *duration_files*, then the attendee at the start of the queue for files immediately starts the files stage, and removes themself from the files queue. If there are no attendees in the queue, then *files_status* is set to free.

On arriving at the clinic, if *files_status* is set to busy, attendees add themselves to the end of the files queue.

The scheduling mechanism works in the same way for the vitals and consultation stages, with attendees adding themselves to the queue for the stage after finishing the files and vitals stages respectively.

##### 1.6.2 Scheduling mechanism – non-rate limiting stages

When the files stage is set to be not rate limiting, then the gap between arrival (the preceding stage) and files, *gap_files*, is kept the same as it is in the attendee input file. Upon arriving at the clinic, each attendee schedules their arrival at files, to occur after a gap of *gap_files*.

The scheduling mechanism works in the same way for the vitals and consultation stages, with the preceding stages being files and vitals respectively.

##### 1.6.3 Attendee waiting areas

In the model, between arrival and files, between files and vitals, and between vitals and consultation, attendees wait in *files_queue_location*, *vitals_queue_location*, and *cons_queue_location* respectively.

###### *1.6.3.1 Queue management intervention*

The exception to this is when the queue management intervention is simulated. The intervention is described more fully below, but briefly, it is assumed in the intervention that a maximum of only $n_1$, $n_2$, and $n_3$ attendees are allowed to wait inside the clinic before each of files, vitals, and consultations respectively, and that the rest wait in a single outdoor waiting area.

Upon arriving the clinic, simulated attendees check how many attendees are currently waiting inside the clinic for the files stage. If it is less than $n_1$, then they wait in *files_queue_location*. If it is greater or equal to $n_1$, then they wait in the outdoor waiting area, and add themselves to the end of a queue.

Each time a attendee reaches files, the length of the queue is checked. If it is greater than zero, then the first attendee in the queue changes their location to *files_queue_location*, and the attendee is removed from the queue.

The process is the same for vitals and consultations.

#### 1.7 Individual characteristics

Individuals in the model are classed as either children (aged <16 years) or adults (aged 16 years or over).

An individual's probability of having pulmonary TB at the time of their clinic visit (*prob_infectious*) is set equal to *prob_infectious_adult* if they are an adult, and *prob_infectious_child* if they are a child.

An individual's breath volume rate ($Ls^{-1}$) (*breathe_rate*) is set equal to *breath_rate_adult* if they are an adult, and *breath_rate _child* if they are a child.

Individuals in the model wear masks with probability *prob_wear_mask*. This is set to zero in the baseline scenario, and in scenarios where no mask wearing intervention is simulated. Each individual has parameters *own_mask_reduction_out* and *own_mask_reduction_in*, which determine any reduction in the rate that they exhale or inhale quanta respectively, that is attributable to the fact they are wearing a mask. They parameters are set to zero if the individual is not wearing a mask, and to *mask_reduction_out* and *mask_reduction_in* respectively if the individual is wearing a mask.

#### 1.8 Room characteristics

Each room has a room volume, *room_volume*, estimated from empirical data[1].

Each room has a rate of air change per hour (ACH), *air_change_rate_h*, which is converted into a rate of air change per time step, *air_change_rate_ts*.

For interventions that had no effect on ventilation rates, the same ventilation rates were used for each run for each paired baseline and intervention model run.

See section 'Intervention scenarios' for details of how *air_change_rate_h* was estimated in intervention scenarios that altered ventilation rates.

The number of adults not wearing masks, children not wearing masks, adults wearing masks, and children wearing masks present in each room were tracked by the parameters *count_adults_no_mask*, *count_children_no_mask*, *count_adults_mask*, and *count_children_mask* respectively.

#### 1.9 Infection risk

Each simulated individual tracks the number of quanta in a room that were produced by themself (*own_quanta_in_room*). This parameter is reset to zero each time an individual changes rooms. Each time step, it is updated using EQ1.

*own_quanta_in_room = (own_quanta_in_room[t-1] * (1 - (air_change_rate_ts)) + prob_infectious * quanta_rate_ts * own_mask_reduction_out)* EQ1

The overall number of quanta in the room over time is tracked using equation EQ2

*quanta_in_room = quanta_in_room[t-1] * (1 - air_change_rate_ts)* EQ2

*+ count_adults_no_mask * prop_infectious_adult * quanta_rate_ts*

*+ count_children_no_mask * prop_infectious_child * quanta_rate_ts*

*+ count_adults_mask * prop_infectious_adult * quanta_rate_ts * mask_reduction_out*

*+ count_children_mask * prop_infectious_child * quanta_rate_ts * mask_reduction_out*

Finally, the risk to each individual each time step is calculated using EQ3

*current_risk* = (1 - exp(- (*quanta_in_room* - *own_quanta_in_room*) * *breath_rate* * *own_mask_reduction_in* / *room_volume*)) EQ3

#### 1.10 Intervention scenarios

##### 1.10.1 Opening windows and doors

Empirical data were available from 20 experiments, where air change rates were estimated in the same room on the same day, both with the doors and windows in a typical in-use configuration ('usual conditions'), and with the doors and windows fully open ('max conditions'). For each of these, the ratio of the air change rate in max conditions compared to usual conditions was estimated. An exponential distribution was fitted to these estimated ratios (Figure 2), and simulated ratios in each room were sampled from the distribution for each model run. Estimated ratios were roughly similar between the data (mean 4.8, median 2.7, IQR 1.4-4.3) and the modelled distribution (mean 3.7, median 2.7, IQR 1.3-5.5).

In the simulated intervention, it is assumed that all doors and windows are kept open at all times.

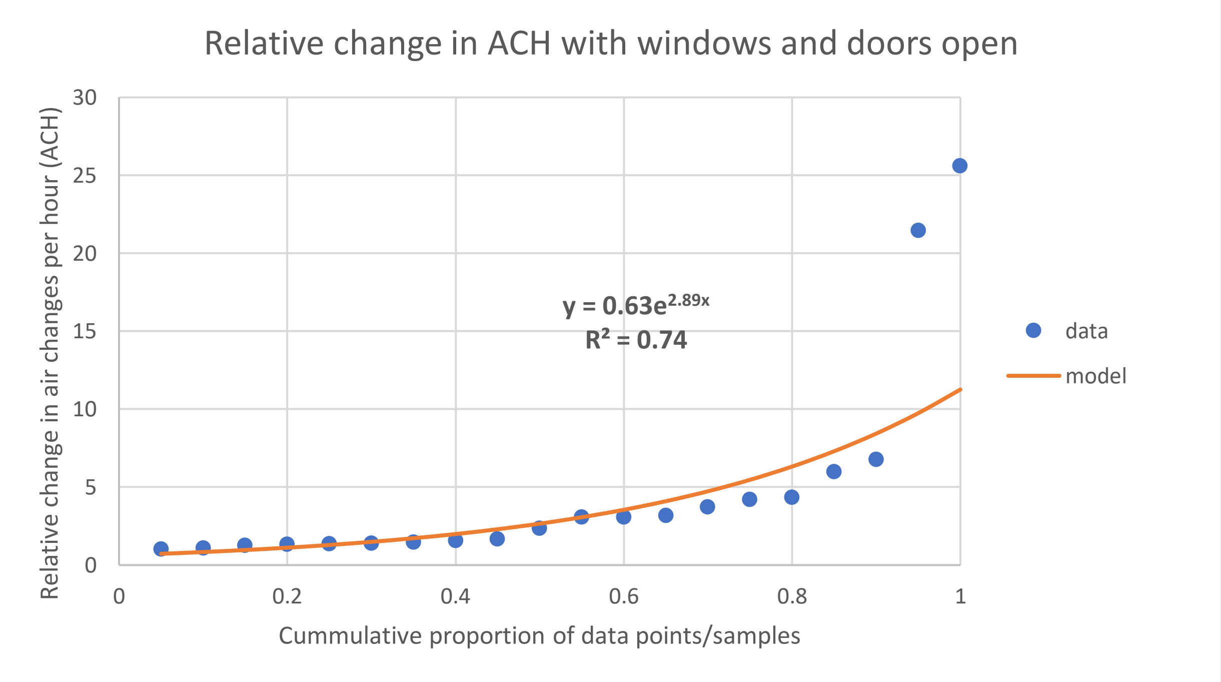

**Figure S2. Empirical data on relative change in air changes per hour (ACH) with doors and windows fully open, compared to windows and doors in their typical configurations; and the distribution used to generate the changes in ACH values in the model**

##### 1.10.2 Simple clinic retrofits

Retrofits are changes to the building to improve ventilation rates. This could include installing lattice brickwork or whirlybird fans. Due to the large amount of variation between clinic spaces in the types of building retrofits that would be suitable, and the lack of sufficient data on the effects of the retrofits on ventilation and air change rates in different types of spaces, we do not model specific retrofits or packages of retrofits. Instead, we simulate an undefined package of retrofits that are sufficient to increase air changes per hour to a minimum of 12 in all rooms, chosen in line with WHO guidelines[7,8]. This is implemented in the model through increasing *air_change_rate_h* to 12 in all rooms and model runs where the sampled air change rate per hour is below 12.

##### 1.10.3 UVGI systems

We assume in this intervention that appropriate and well maintained ultraviolet germicidal irradiation (UVGI) systems are installed in all indoor clinic waiting areas.

Empirical data from studies of transmission to guinea pigs suggest that UVGI reduces the rate of transmission by 80% (95% CI 64%-88%)[9], equivalent to a ventilation rate of 24 ACH (95% CI 9.9-62)[9]. This is implemented in the model through an additional quanta clearance rate, simulated in the same way as clearance through ventilation. The value of the additional quanta clearance rate is sampled for each waiting area and model run from a split normal distribution with mean 24 and 95% CI 9.9-62%.

##### 1.10.4 Surgical masks wearing by clinic attendees

Based on discussions with health care workers and professionals active in the management of health services in the two provinces we worked in, as well as review of qualitative data collected, we determined that a scenario where 70% of attendees wear surgical masks 90% of the time was plausible. This is implemented in the model as 63% of attendees wearing masks 100% of the time, with the attendees who wear the masks chosen at random each model run.

The relative reduction in the quanta production rate for each mask-wearing attendee each run is assumed to be the same, and the reduction is sampled for each model run from a split normal distribution with mean 75% and 95% CI 56-85%[10].

We assume that masks have no effect on risk of infection for the person wearing the mask[11].

##### 1.10.5 Increased CCMDD coverage

South Africa's Central Chronic Medicine Dispensing and Distribution (CCMDD) programme is designed to allow patients with stable chronic health conditions to collect their medicines from convenient locations, such as local pharmacies[12]. This means that they do not need to queue at clinics unnecessarily. The purpose of this intervention is to increase the coverage of CCMDD and similar programmes for eligible patients on ART, and to ensure that pick-up points do not require patients to queue at clinics.

In simulating the intervention, we focus on ART patients only, as they make up a large proportion of patients attending for non-tuberculosis related chronic care (399/493, 81%, in the empirical datasets), and because few data were available on patients with other chronic conditions such as diabetes.

Visit reason was collected from all attendees visiting the clinics on the data collection days, with 'Chronic care: HIV/ART' being one of the reported reasons. We assume that some of the clinic visits with 'Chronic care: HIV/ART' being listed as the main visit reason would not be needed with the increased implementation of CCMDD. We therefore remove a proportion, *p*, of those attendees from the model.

In Western Cape clinics, there was an error during data collection, with the majority of patients who attended for HIV/ART related reasons having their main visit reason recorded as 'Acute care: minor problems'[13]. The correct proportions of adult male and female patients attending for HIV/ART related reasons were therefore estimated for Western Cape clinics from the proportions in the

KwaZulu-Natal clinics, adjusted for the lower prevalence of HIV and ART coverage in Western Cape[14]. Adult Western Cape 'Acute care: minor problems' patients were then assigned at random, for each clinic and model run, to have attended for HIV/ART related reasons, to reach the desired proportion of male and female patients attending for HIV/ART related care.

2.8% of attendees report their visit reason as attending on behalf of somebody else. We assume that a proportion, *p*, of those visits would also need not occur under a scaled up CCMDD intervention.

For 69/120 (59%) people who reported their visit reason as accompanying an adult, and 72/179 (40%) people who reported their visit reason as accompanying a child, the visit reason of the person that they were accompanying could be determined. For accompanying people for whom the visit reason of the person they were accompanying could not be determined, they were randomly assigned to be accompanying an HIV/ART patient each model run, with probability equal to the proportions where it could be determined, by clinic and whether they were accompanying an adult or a child. A proportion, *p*, of the visits of people assigned to accompanying someone attending the clinic for HIV/ART care were assumed not to have been needed under the intervention scenario.

The proportion, *p*, was determined using data from a social contacts survey of 1704 adults living in the catchment areas of two clinics in KwaZulu-Natal[15]. Respondents were asked to report the number of times that they had attended a clinic (for their own health) in the past six months. Self-reported HIV-positive people (of who 480/493 (97%) reported being on ART), reported a mean of 8.8 clinic visits per year, compared to 4.1 by HIV-/unknown. That is, an excess of 4.1 (95% CI 3.6-4.5) visits per year, controlling for age and sex, which we attribute to ART appointments. We assume that 92% (95% CI 84-95%) of people could have their ART appointments reduced to once every 6 months (the estimated proportion of people on ART who were virally suppressed 2019[14]), and that the remaining 8% of people need monthly ART appointments. This gives us a 31% reduction (IQR 22-34%) in HIV/ART care visits. For each clinic and model run, the number of excess visits and proportion of ART patients who are virally supressed are sampled from the relevant normal distributions, and *p* is calculated.

We implicitly assume that CCMDD pickup either occurs at a location away from the clinic; or requires patients to spend a negligible amount of time inside the clinic, without having any effect on the delays for other patients.

Ethical approval for the social contacts survey was granted by the Biomedical Research Ethics Committee (REC) of the University of KwaZulu-Natal (UKZN) (BE662/17) and the London School of Hygiene & Tropical Medicine (14640).

##### 1.10.6 Queue management system and outside waiting areas

Empirical data show that clinic waiting areas are often crowded, and that in many clinics attendees wait in unsuitable areas such as corridors[13]. Conversations with clinic staff suggested that this is partly due to patient concerns that if they wait in other areas, they may not hear their name being called, and may miss their turn. This intervention therefore combines a large, covered outdoor waiting area with a queue management system, such as numbered tickets or an electronic tracking system.

We assume in the model that only the next $n_1$, $n_2$, and $n_3$ attendees due to be seen at files, vitals, or for consultations respectively are allowed to wait inside the clinic. At smaller clinics, with fewer than 300 attendees on the day of data collection, $n_1$=5, $n_2$=5, and $n_3$=10. At larger clinics, $n_1$=10, $n_2$=10, and $n_3$=20. Once allowed inside the clinic, attendees are assumed to wait in the same location for each stage as they wait in the baseline scenario.

The volume of the outdoor waiting area is assumed to be equal to the sum of the volume of the existing clinic waiting areas. The ACH is the outdoor waiting area is drawn from a uniform distribution between 52 and 70 ACH for each clinic and model run[16]

##### 1.10.7 Appointment systems

In this intervention, we simulate an appointment system to reduce clinic overcrowding, through spacing out the arrival times of patients. As date-time appointment systems were already in place in some form in the Western Cape clinics on the day that the attendee data were collected, we only model the appointment intervention in the KwaZulu-Natal clinics.

We assume that appointments are given in 10-minute slots (i.e. a patient could be assigned 10:00 or 10:10, but not 10:05), between 9am and 1.50pm, and that patients arrive between 0-10 minutes before their appointment (sampled from a uniform distribution for each attendee). Once arrived at the clinic, simulated attendees are seen by clinic staff as soon as capacity allows, even if it is before their appointment time. Arrival times are not changed for attendees who are not assigned appointments, and they enter the simulated queues at the time that they arrive at the clinic.

Patients were assumed to be acute patients if their main reported visit reason was ‘Acute care: minor problems’ or ‘Acute care: 24-hour emergency unit’, and chronic otherwise. As with the CCMDD intervention, a proportion of patients at Western Cape clinics whose visit reason was recorded as ‘Acute care: minor problems’ was assumed to have visited for HIV/ART care – i.e. chronic care. In the model, appointments are given to all adult chronic patients. The first N acute patients are assumed to be seen the same day, as well as any children aged <16 years. The remaining adult acute patients are given appointments.

N is calculated for each clinic and model run by multiplying the total number of attendees counted on the day of data collection by the proportion of the total daily clinic time (length of time set aside for drop-in acute patients only plus the length of time that the clinic assigns appointments) that is set aside to see patients without appointments in the morning. N is then multiplied by a number drawn from a random uniform distribution between 0.75 and 1.25 for each clinic and model run, to reflect day-to-day fluctuations in the numbers of patients.

For 69/120 (59%) people who reported their visit reason as accompanying an adult, and 72/179 (40%) people who reported their visit reason as accompanying a child, the visit reason of the person that they were accompanying could be determined. For accompanying people for whom the visit reason of the person they were accompanying could not be determined, they were randomly assigned to be accompanying an acute or chronic patient each model run, with probability equal to the proportions where it could be determined, by clinic and accompanying adult or child. Accompanying people were given appointments or seen the same day based on the visit reason of the person they were accompanying.

It is assumed that there is no risk of transmission to or from attendees while they are receiving their appointment slots, reflecting the fact that many appointments could be arranged on a prior visit or by telephone, and that the remaining appointments could be arranged quickly in a well ventilated or covered outdoor location, with the attendees rapidly leaving the clinic after receiving their appointment.

In the appointment system intervention, when the files stage in considered to be rate limiting (see section 'Movement through clinics'), the gap between attendees at files is reduced by 50%. This is done to incorporate a plausible reduction in the mean time taken to find files that might be achieved by pre-retrieval of files for patients with appointments.

#### 1.11 Input parameter values

| Parameter | Scenario | Description | Value | Source |
|---|---|---|---|---|
| *prob_infectious_adult* | All | Proportion of adults visiting the clinic that have pulmonary TB | 0.010 | Clinic prevalence survey[5] |
| *prob_infectious_child* | All | Proportion of children visiting the clinic that have pulmonary TB | 0.00016 | Clinic prevalence survey[5], adjusting for lower proportion of smear+ disease in children[3], and lower incidence of disease[4] |
| *quanta_rate_hour* | All | Rate of quanta production per hour for individuals with pulmonary TB | 1.25 | Andrews *et al* (2014)[6] |
| *breath_rate_adult* | All | Breath volume rate of adults ($lh^{-1}$) | 480 | Rudnick and Milton (2003)[17] |
| *breath_rate_child* | All | Breath volume rate of children ($lh^{-1}$) | 288 | Rudnick and Milton (2003)[17], adjusting for lower breathe volume in children[18] |

| | | | | |
|---|---|---|---|---|
| *quanta_rate_ts* | All | Time step for updating quanta and infection risk estimates (seconds) | 10 | NA |
| *min_ACH* | Retrofits | Minimum air changes per hour | 12 | WHO guidelines[7,8] |
| *mask_reduction_out* | Masks | Relative rate of quanta exhalation in individuals with pulmonary TB who wear are mask compared to those who don't | 0.25 (0.15-0.44) | Dharmadhikari *et al* (2012)[10] |
| *mask_reduction_in* | Masks | Relative rate of quanta inhalation in individuals with pulmonary TB who wear are mask compared to those who don't | 1 | MacIntyre (2015)[11] |
| *prob_wear_mask* | Masks | Proportion of attendees who wear a surgical mask | 0.9 * 0.7 = 0.63 | Expert opinion |
| *UVGI_rate* | UVGI | Rate of quanta clearance due to UVGI, given in units of the equivalent air changes per hour | 24 ACH (95% CI 9.9-61.7) | Mphaphlele (2015)[9] |

| | | | | |
|---|---|---|---|---|
| *files_indoor_number* | Queue management | Number of attendees allowed to wait inside the clinic for the files step | Clinics 2, 8, 9, 12: 5; Clinics 1, 5, 6, 11: 10 | Expert opinion |
| *vitals_indoor_number* | Queue management | Number of attendees allowed to wait inside the clinic for the files step | Clinics 2, 8, 9, 12: 5; Clinics 1, 5, 6, 11: 10 | Expert opinion |
| *consultation_indoor_number* | Queue management | Number of attendees allowed to wait inside the clinic for the files step | Clinics 2, 8, 9, 12: 5; Clinics 1, 5, 6, 11: 10 | Expert opinion |
| *outdoor_waiting_area_ACH* | Queue management | Air changes per hour (ACH) in the outdoor waiting area | 52-70 | Escombe *et al*[16] |
| *excess_visits_ART* | CCMDD | Number of excess clinic visits per year for people on ART, compared to people not on ART | 4.1 (95% CI 3.6-4.5) | Empirical social contact data[15] |
| *prop_viral_supressed* | CCMDD | Proportion of patients on ART who are virally supressed | 92% (95% CI 84-95%) | AIDSinfo[19] |

### 2 Supplementary results

#### 2.1 Sensitivity analysis

Simulating consultations as a non-rate limiting stage reduced the estimated reduction in the rate of transmission to 15% (IQR 8.7-23%) in the CCMDD scale-up intervention, and 24% (IQR 13-47%) in the appointments intervention (Figure S3). It had no effects on the estimates for any other intervention.

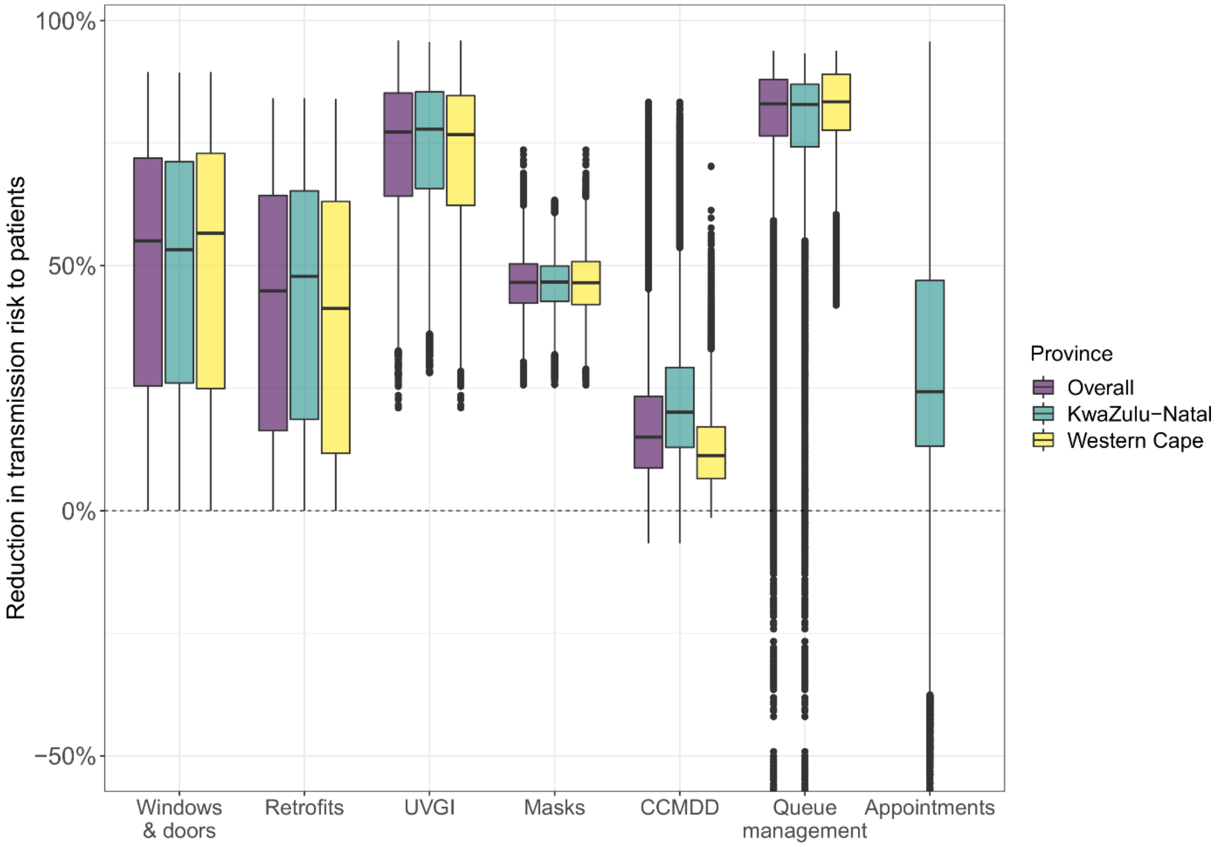

**Figure S3. Estimated reduction in the rate of *Mycobacterium tuberculosis* transmission to attendees in clinics, by province and intervention, when consultations are included in the model as a non-rate limiting stage.** The central line indicates the median, the box range the interquartile range (IQR), the whiskers the most extreme value within 1.5 * IQR from the box, and the points outlying values. In the queue management intervention in KwaZulu-Natal, 1% of points were below -50%, with a minimum of -162%. In the appointments intervention in KwaZulu-Natal, 0.28% of points were below -50%, with a minimum of -150%. These points are not shown on the graph. The appointment system intervention was not modelling in Western Cape, due to the presence of existing appointment systems. UVGI stands for ultraviolet germicidal irradiation, and CCMDD for Central Chronic Medicine Dispensing and Distribution.

#### 2.2 Intervention impact by clinic

Figure S4 shows the effect of the interventions on the rate of transmission to attendees by clinic.

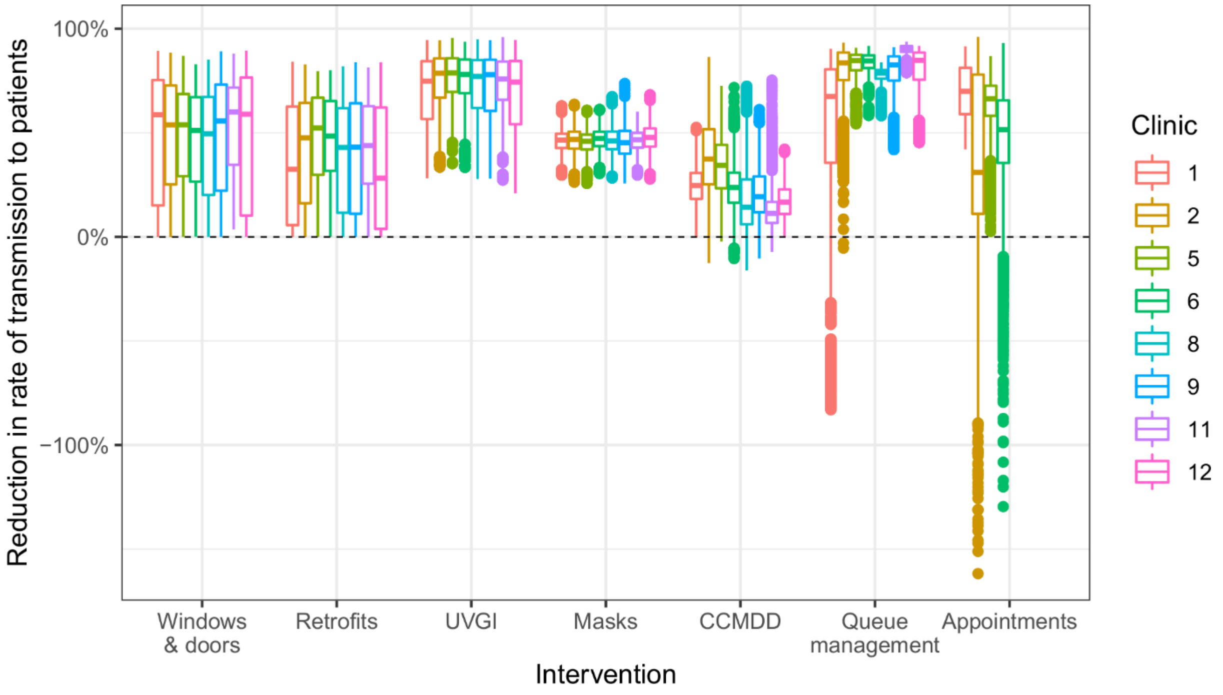

**Figure S4. Estimated reduction in the rate of *Mycobacterium tuberculosis* transmission to attendees in clinics, by clinic and intervention.** The central line indicates the median, the box range the interquartile range (IQR), the whiskers the most extreme value within 1.5 * IQR from the box, and the points outlying values. The appointment system intervention was not modelling in Western Cape, due to the presence of existing appointment systems. UVGI stands for ultraviolet germicidal irradiation, and CCMDD for Central Chronic Medicine Dispensing and Distribution.

#### 2.3 Attendee numbers and rate of transmission over time

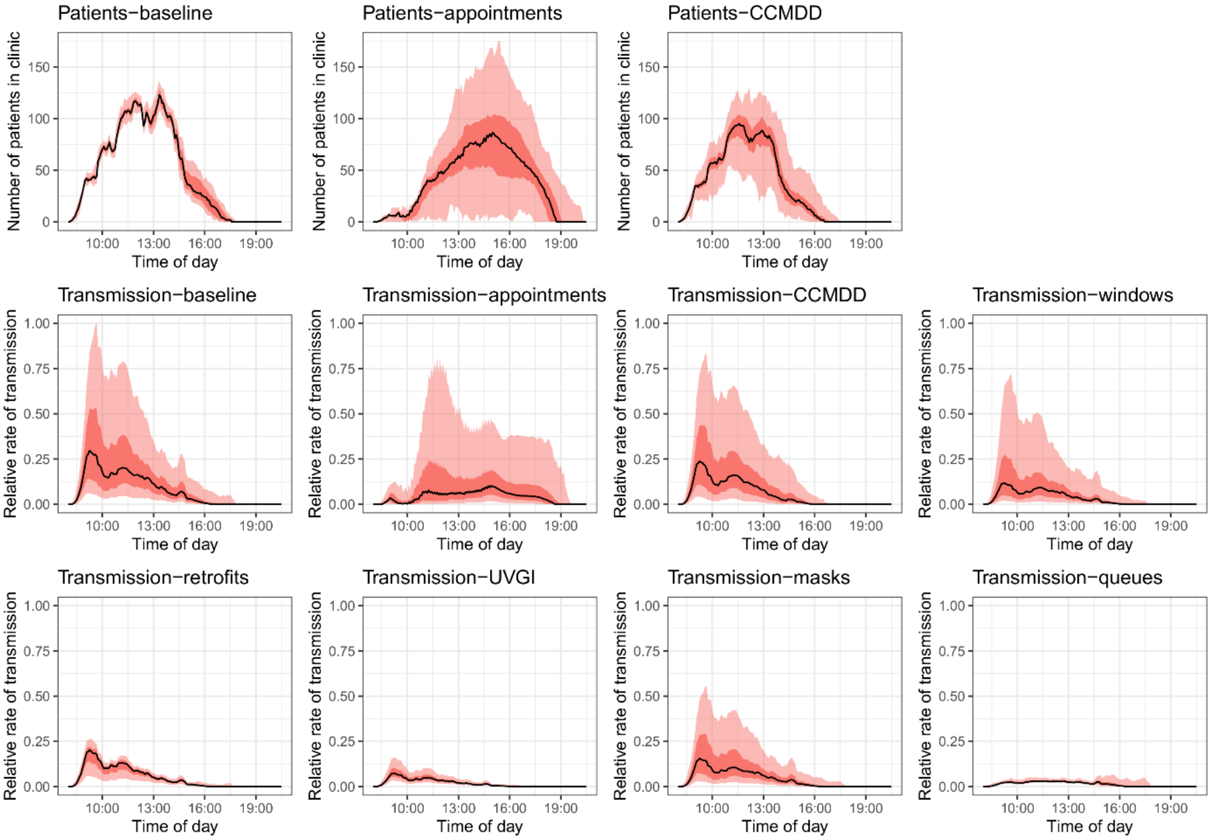

**Figure S5. Number of attendees in the clinic over time in the baseline, appointments, and CCMDD interventions, and the mean rate of transmission to each attendee in the clinic over time in all scenarios, for clinic 2.** The black line shows the median result, the dark red band the interquartile range, and the light red band the 95% plausible range. For interventions where a plot of the number of attendees over time is not shown, the intervention has no effect on attendee numbers. Transmission rates are relative to the highest transmission rate in any scenario at any point in time. UVGI stands for ultraviolet germicidal irradiation, and CCMDD for Central Chronic Medicine

Dispensing and Distribution.

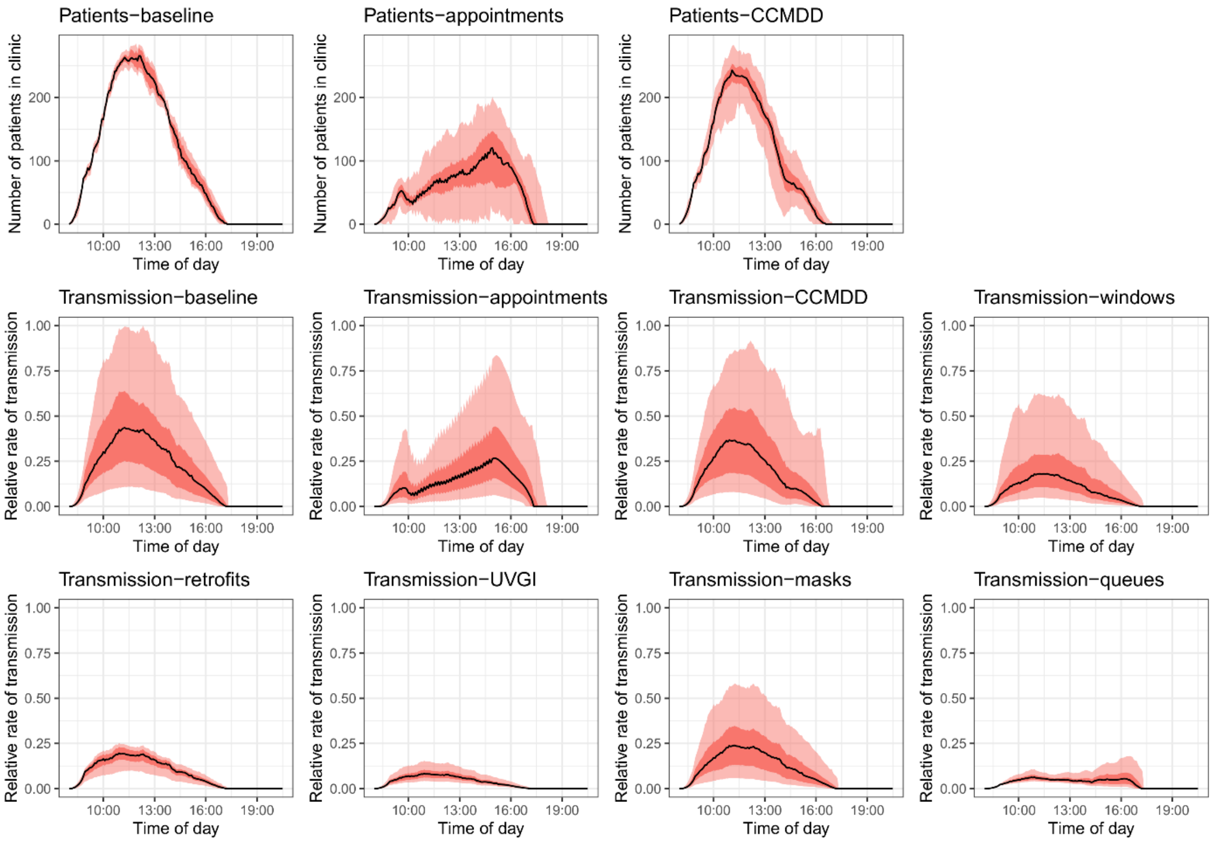

**Figure S6. Number of attendees in the clinic over time in the baseline, appointments, and CCMDD interventions, and the mean rate of transmission to each attendee in the clinic over time in all scenarios, for clinic 5.** The black line shows the median result, the dark red band the interquartile range, and the light red band the 95% plausible range. For interventions where a plot of the number of attendees over time is not shown, the intervention has no effect on attendee numbers. Transmission rates are relative to the highest transmission rate in any scenario at any point in time. UVGI stands for ultraviolet germicidal irradiation, and CCMDD for Central Chronic Medicine Dispensing and Distribution.

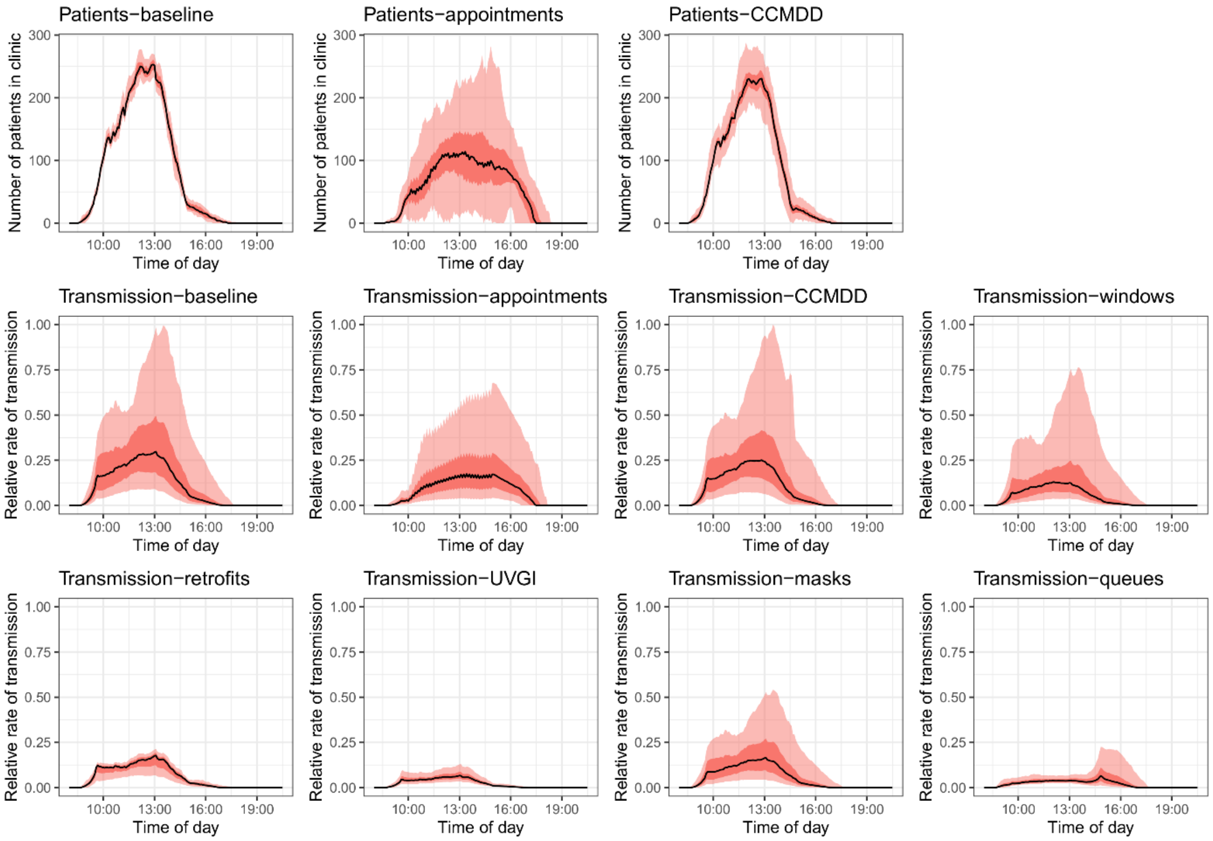

**Figure S7. Number of attendees in the clinic over time in the baseline, appointments, and CCMDD interventions, and the mean rate of transmission to each attendee in the clinic over time in all scenarios, for clinic 6.** The black line shows the median result, the dark red band the interquartile range, and the light red band the 95% plausible range. For interventions where a plot of the number of attendees over time is not shown, the intervention has no effect on attendee numbers. Transmission rates are relative to the highest transmission rate in any scenario at any point in time. UVGI stands for ultraviolet germicidal irradiation, and CCMDD for Central Chronic Medicine Dispensing and Distribution.

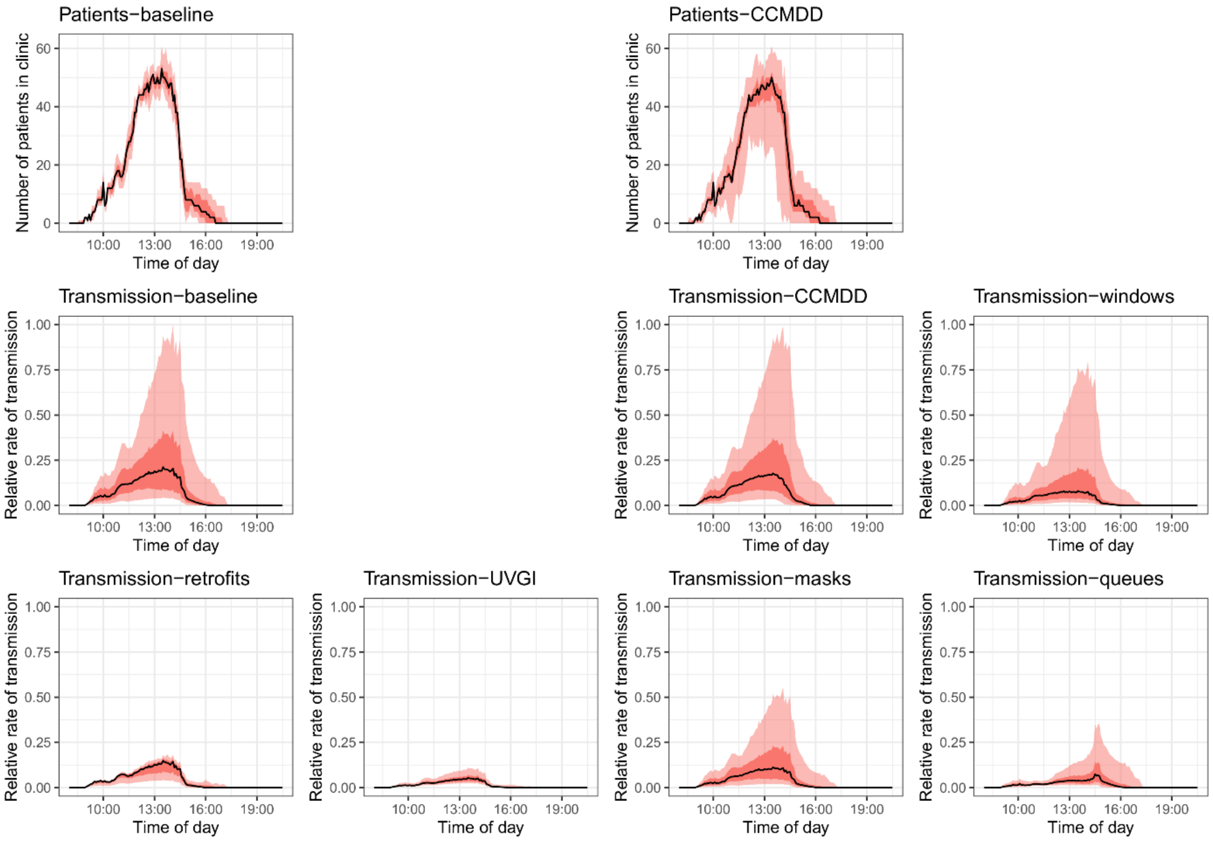

**Figure S8. Number of attendees in the clinic over time in the baseline, appointments, and CCMDD interventions, and the mean rate of transmission to each attendee in the clinic over time in all scenarios, for clinic 8.** The black line shows the median result, the dark red band the interquartile range, and the light red band the 95% plausible range. For interventions where a plot of the number of attendees over time is not shown, the intervention has no effect on attendee numbers. Transmission rates are relative to the highest transmission rate in any scenario at any point in time. UVGI stands for ultraviolet germicidal irradiation, and CCMDD for Central Chronic Medicine Dispensing and Distribution.

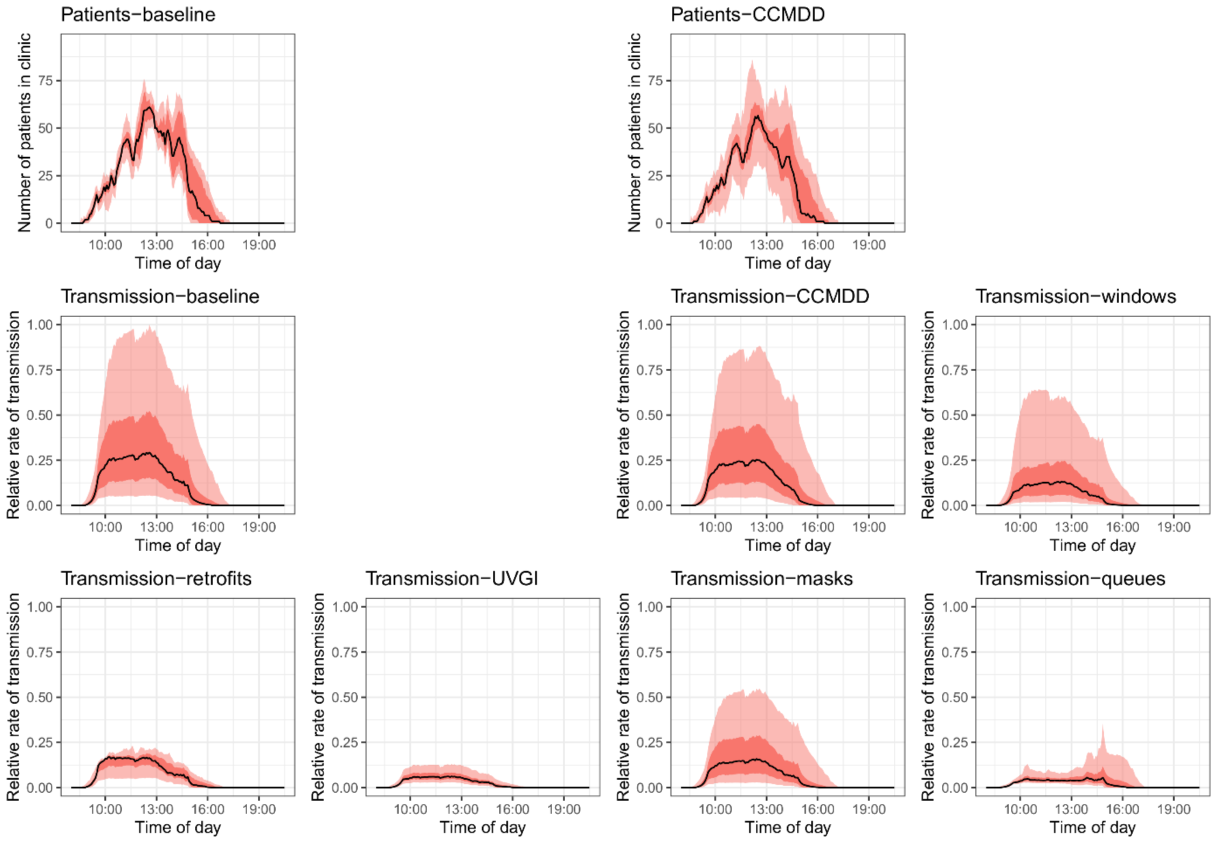

**Figure S9. Number of attendees in the clinic over time in the baseline, appointments, and CCMDD interventions, and the mean rate of transmission to each attendee in the clinic over time in all scenarios, for clinic 9.** The black line shows the median result, the dark red band the interquartile range, and the light red band the 95% plausible range. For interventions where a plot of the number of attendees over time is not shown, the intervention has no effect on patient numbers. Transmission rates are relative to the highest transmission rate in any scenario at any point in time. UVGI stands for ultraviolet germicidal irradiation, and CCMDD for Central Chronic Medicine Dispensing and Distribution.

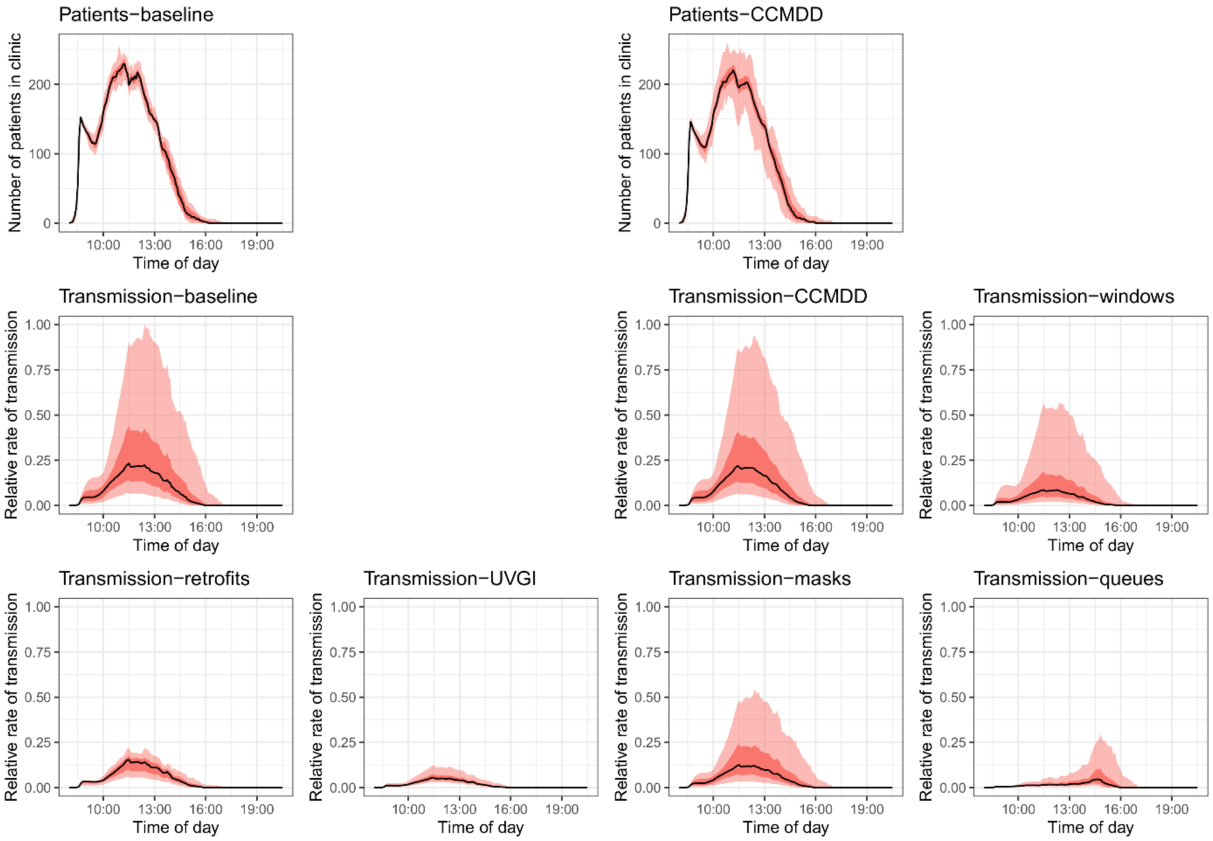

**Figure S10. Number of attendees in the clinic over time in the baseline, appointments, and CCMDD interventions, and the mean rate of transmission to each attendee in the clinic over time in all scenarios, for clinic 11.** The black line shows the median result, the dark red band the interquartile range, and the light red band the 95% plausible range. For interventions where a plot of the number of attendees over time is not shown, the intervention has no effect on attendee numbers. Transmission rates are relative to the highest transmission rate in any scenario at any point in time. UVGI stands for ultraviolet germicidal irradiation, and CCMDD for Central Chronic Medicine Dispensing and Distribution.

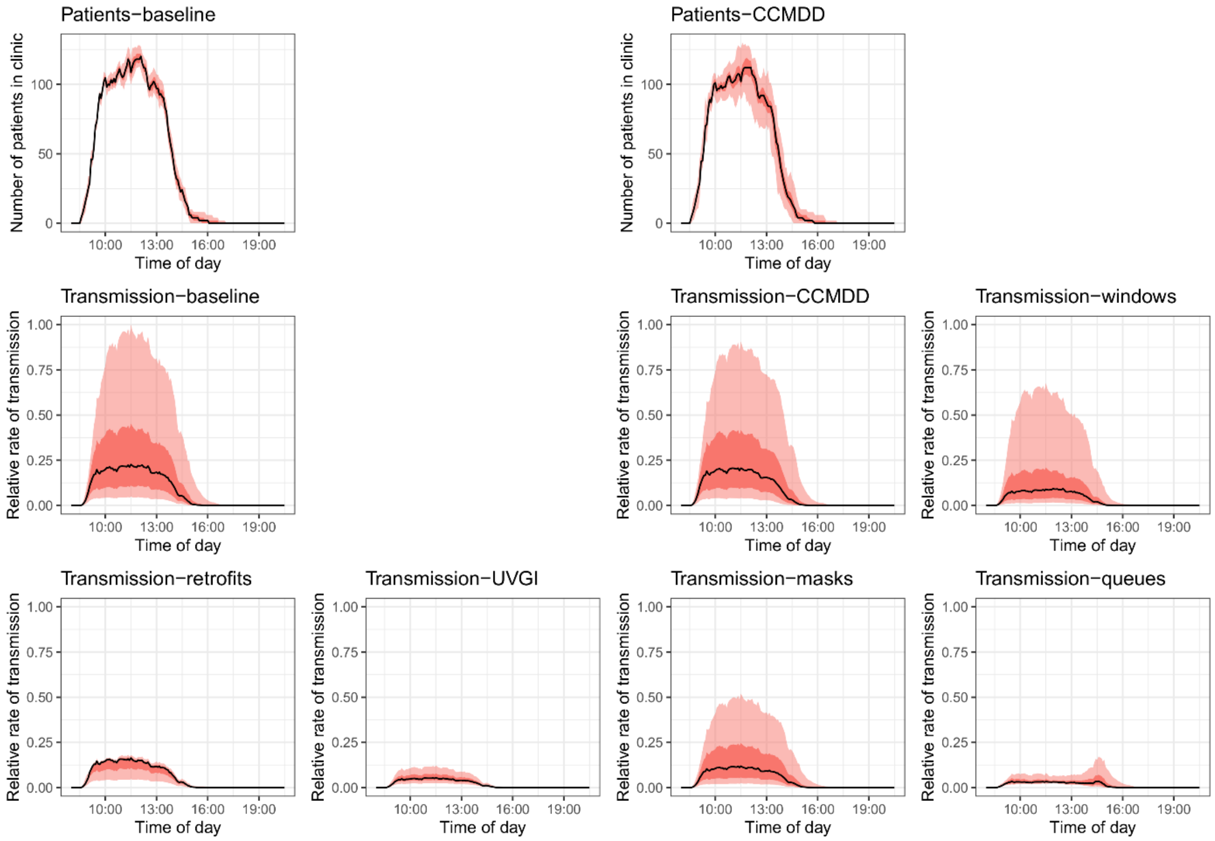

**Figure S11. Number of attendees in the clinic over time in the baseline, appointments, and CCMDD interventions, and the mean rate of transmission to each attendee in the clinic over time in all scenarios, for clinic 12.** The black line shows the median result, the dark red band the interquartile range, and the light red band the 95% plausible range. For interventions where a plot of the number of attendees over time is not shown, the intervention has no effect on attendee numbers. Transmission rates are relative to the highest transmission rate in any scenario at any point in time. UVGI stands for ultraviolet germicidal irradiation, and CCMDD for Central Chronic Medicine Dispensing and Distribution.

#### 2.4 Median clinic visit durations

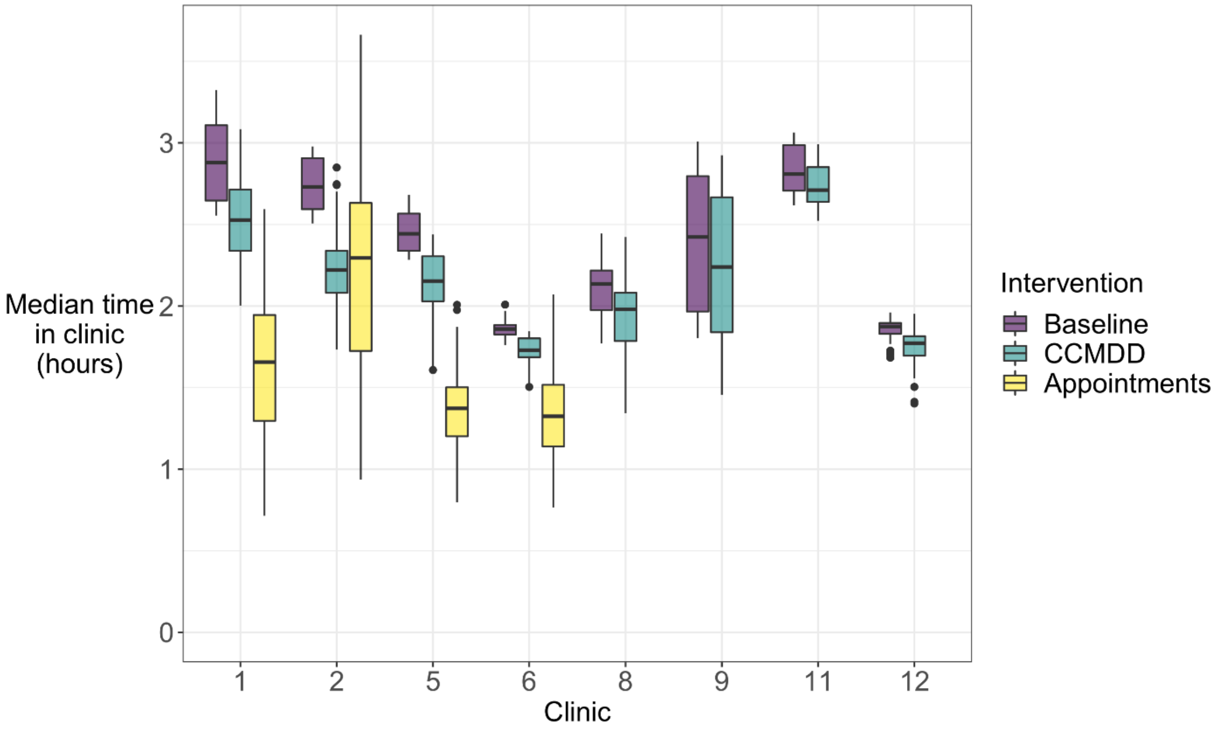

**Figure S12. Median attendee times in clinic, by clinic and intervention.** The boxplots show the distribution of the median attendee time in the clinic for each model run (i.e. not the duration of time spent in the clinic by each attendee). The central line indicates the median across model runs, the box ranges the interquartile range (IQR), the whiskers the most extreme value within 1.5 * IQR from the box, and the points outlying values. CCMDD for Central Chronic Medicine Dispensing and Distribution.

### 3 Supplementary acknowledgements

*The extended Umoya omuhle team, institutions, and roles (listed alphabetically by surname)*

| Name | Institution/s | Role |
|---|---|---|
| Siphokazi Adonisi | UCT | Research Assistant |
| Kathy Baisley | LSHTM; AHRI | Co-investigator |
| Peter Beckwith | LSHTM; UCT | Research fellow |
| Fiammetta Bozzani | LSHTM | Co-investigator |
| Amy Burdzik | UCT | Occupational health |
| Adrienne Burrough | LSHTM | Project Manager |
| Nkosingiphile Buthelezi | AHRI | Research Assistant |
| Xolile Buthelezi | AHRI | Diagnostic Lab Manager |
| Ruvimbo Chigwanda | UCT | Administration |

| Name | Institution/s | Role |
|---|---|---|
| Christopher Colvin | UCT | Co-investigator |
| PIP CRAs | AHRI | Clinic research Assistants |
| Njabulo Dayi | AHRI | Research Data Manager |
| Arminder Deol | LSHTM | Mathematical modeller |
| Karina Diaconu | QMU | Co-investigator |
| Siphephelo Dlamini | AHRI | Nursing Manager |
| Yutu Dlamini | AHRI | Research Assistant |
| Raveshni Durgiah | AHRI | Grants office |
| Anita Edwards | AHRI | Head: Scientific Support |
| Jennifer Falconer | QMU | Research Assistant |
| Kitty Flynn | QMU | Administrator |
| Patrick Gabela | AHRI | Clinical Research Data Coordinator |
| Dickman Gareta | AHRI | Head: Research Data Management |
| Awethu Gawulekapa | UCT | Research Assistant |
| Harriet Gliddon | AHRI; UCL | Research Assistant |
| Bavashni Govender | UKZN | Administration |
| Indira Govender | LSHTM; AHRI | Co-investigator |
| Alison Grant | LSHTM; AHRI | Principal investigator |
| Meghann Gregg | LSE | Research fellow |
| Emmerencia Gumede | AHRI | Research Assistant |
| Sashin Harilall | AHRI | Grants office |
| Kobus Herbst | AHRI | Chief Information Officer |
| Tamia Jansen | UCT | Research Assistant |
| Seonaid Kabiah | UCT | Research Assistant |
| Idriss Kallon | UCT | Post-doctoral researcher |
| Aaron Karat | LSHTM | Co-investigator |
| Hannah Keal | AHRI | Communications |
| Suzanne Key | UCT | Occupational health |
| Zama Khanyile | UKZN | Research Assistant |
| Mandla Khoza | AHRI | Clinic Research Assistant |
| Nozi Khumalo | AHRI | Systems Engineer |
| Zilethile Khumalo | AHRI | Research Assistant |
| Karina Kielmann | QMU | Co-principal investigator |
| Nondumiso Kumalo | AHRI | Clinic Research Assistant |
| Richard Lessells | AHRI | Epidemiologist |
| Nokuthula Lushaba (deceased) | UKZN | Administration |
| Sithembiso Luthuli | AHRI | Research Assistant |
| Sinethemba Mabuyakhulu | AHRI | Clinic Research Assistant |
| Hayley MacGregor | IDS | Co-investigator |
| Nonhlanhla Madlopha | AHRI | Research Assistant |
| Aphiwe Makalima | UCT | Administration |
| Tacha Malaza | AHRI | PIP CRA |
| Sifundesihle Malembe | AHRI | Research Assistant |
| Godfrey Manuel | UCT | Transport |

| Name | Institution/s | Role |
|---|---|---|
| Nonhlanhla Maphumulo | UKZN | Administration |
| Precious Mathenjwa | UCT | Research Assistant |
| Sanele Mbuyazi | AHRI | PIP CRA |
| Nicky McCreesh | LSHTM | Co-investigator |
| Claire McLellan | QMU | Administrator |
| Simphiwe Mdluli | AHRI | PIP CRA |
| Thabile Mkhize | AHRI | Transport |
| Duduzile Mkhwanazi | AHRI | Research Assistant |
| Zinhle Mkhwanazi | AHRI | Research Assistant |
| Zodwa Mkhwanazi | AHRI | Research Assistant |
| Anathi Mngxekeza | UCT | Research Assistant |
| Tshwaraganang Modise | AHRI | Research Data |
| Sashen Moodley | AHRI | Microbiology Laboratory Supervisor |
| Samantha Moyo | UCT | Research Assistant |
| Silindile Mthembu | AHRI | Clinic Research Assistant |
| Nozipho Mthethwa | AHRI | Research Assistant |
| Siphesihle Mthethwa | AHRI | Procurement Coordinator |
| Sphiwe Mthethwa | AHRI | Research Assistant |
| Sanele Mthiyane | AHRI | Research Assistant |
| Vanisha Munsamy | AHRI | Grants office |
| Sinead Murphy | UCT | Research Assistant |
| Thomas Murray | AHRI | Research assistant |
| Senzile Myeni | AHRI | PIP CRA |
| Tevania Naidoo | AHRI | Procurement |
| Nompilo Ndlela | AHRI | Research Assistant |
| Zama Ndlela | AHRI | PIP CRA |
| Thandekile Nene | AHRI | Research Assistant |
| Phumla Ngcobo | AHRI | Communications |
| Nzuzo Ntombela | AHRI | Research Data Systems Service Manager |
| Sabelo Ntuli | AHRI | GIS Coordinator |
| Nompumulelo Nyawo | AHRI | Human resources |
| Phumzile Nywagi | UCT | Research Assistant |
| Stephen Olivier | AHRI | Statistician |
| Justin Parkhurst | LSE | Co-investigator |
| Alex Pym | AHRI | Co-investigator |
| Yolanda Qeja | UCT | Research Assistant |
| Anand Ramnanan (deceased) | AHRI | Procurement |
| Sharmila Rugbeer | UKZN | Administration |
| Janet Seeley | LSHTM | Co-investigator |
| Aruna Sevakram | AHRI | Scientific support |
| Sizwe Sikhakane | AHRI | Transport |
| Zizile Sikhosana | AHRI | Somkhele Laboratory Supervisor |
| Theresa Smit | AHRI | Head: Diagnostic Research |
| Thandeka Smith | UKZN | Research Assistant |

| Name | Institution/s | Role |
|---|---|---|
| Naomi Stewart | LSHTM | Communications |
| Alison Swartz | UCT | Co-investigator |
| Amy Thomas | LSHTM | Communications |
| Siphosethu Titise | UCT | Research Assistant |
| Anna Vassall | LSHTM | Co-investigator |
| Marlise Venter | AHRI | Facilities Administrator |
| Anna Voce | UKZN | Co-investigator |
| Richard White | LSHTM | Co-investigator |
| Tom Yates | Imperial | Co-investigator |
| Precious Zulu | AHRI | Administration |
| Gimenne Zwama | QMU | Research Fellow |

AHRI: Africa Health Research Institute; IDS: Institute of Development Studies; LSE: London School of Economics and Political Science; LSHTM: London School of Hygiene & Tropical Medicine; QMU: Queen Margaret University; UCT: University of Cape Town; UKZN: University of KwaZulu-Natal;
